# Levels and Determinants of Person-Centered Maternity Care Among Women Living in Urban Informal Settlements: Evidence from Client Exit Surveys in Nairobi, Lusaka and Ouagadougou

**DOI:** 10.1101/2025.01.09.25320278

**Authors:** Safia S. Jiwani, Kadari Cisse, Martin Mutua, Choolwe Jacobs, Anne Njeri, Godfrey Adero, Mwiche Musukuma, Dennis Ngosam, Fatou Sissoko, Seni Kouanda, Amanuel Abajobir, Cheikh Faye, Ties Boerma, Agbessi Amouzou

**Author notes:** Corresponding author: Safia S. Jiwani, Department of International Health, Johns Hopkins Bloomberg School of Public Health; 615 N Wolfe St, Baltimore MD 21205.

## Abstract

**Background:** Sub-Saharan Africa’s rapid urbanization has led to the sprawling of urban informal settlements. The urban poorest women are more likely to experience worse health outcomes and poor treatment during childbirth. This study measures levels of person-centered maternity care (PCMC) and identifies determinants of PCMC among women living in urban informal settlements in Nairobi, Lusaka and Ouagadougou.

**Methods:** We conducted phone, home-based or facility-based exit surveys of women discharged from childbirth care in facilities serving urban informal settlements. We estimated overall and domain-specific PCMC scores covering dignity and respect, communication and autonomy, and supportive care. We ran multilevel linear regression models to identify structural, intermediary and health systems factors associated with PCMC.

**Results:** We included 1,249 women discharged from childbirth care: the majority were aged 20-34 years and were unemployed. In Lusaka and Nairobi, over 65% of women had secondary education, and over half gave birth in a hospital, whereas in Ouagadougou a third had secondary education and 30.4% gave birth in a hospital. The mean PCMC score ranged from 57.1% in Lusaka to 73.8% in Ouagadougou. Across cities, women reported high dignity and respect mean scores (73.5% -84.3%), whereas communication and autonomy mean scores were consistently poor (47.6% - 63.2%). In Ouagadougou, women with formal employment, those who delivered in a private for-profit facility, and whose newborn received postnatal care before discharge reported significantly higher PCMC. In Nairobi and Lusaka, women who were attended by a physician during childbirth, and those whose newborn was checked before discharge reported significantly higher PCMC.

**Conclusion:** Women living in urban informal settlements experience inadequate PCMC and report poor communication with health providers. Select health systems and provision of care factors are associated with PCMC in this context. Quality improvement efforts are needed to enhance PCMC and ensure women’s continuity in care seeking.

**Key Messages:** *What is already known on this topic:* Despite high use of maternal and newborn health services in urban areas, health outcomes still remain worse among lower-income populations, and we know little about the quality of services and experience of care among the urban poorest women. Studies suggest that women who experience disrespect and abuse during childbirth are more likely to discontinue using health services. Person-centered maternity care (PCMC) refers to care that is respectful of and responsive to women’s needs, preferences and values. Previous studies have reported sub-optimal levels of person-centered maternity care in low-and middle-income settings. We conducted this study to evaluate the levels of PCMC and identify structural, intermediary and health systems factors associated with PCMC among low-income urban women living in informal settlements in sub-Saharan African capital cities.

*What this study adds:* Women living in urban informal settlements in Nairobi, Lusaka and Ouagadougou experience inadequate PCMC, with overall mean scores ranging from 57.1% (51.4 points out of 90) to 73.8% (66.4 points out of 90). Most women reported experiencing dignity and respect during childbirth, but communication with providers was consistently poor, with mean scores ranging from 47.6% (12.8 points out of 27) in Lusaka to 63.2% (17.1 points out of 27) in Nairobi. In Ouagadougou, women with formal employment, those who delivered in a private for-profit facility, and whose newborn received postnatal care prior to discharge reported significantly higher PCMC. In Nairobi and Lusaka, women who were attended by a physician during childbirth, and those whose newborn received postnatal care before discharge reported significantly higher PCMC.

*How this study affects research, practice or policy:* Further research is needed to understand health providers’ barriers in offering PCMC and the structures enabling PCMC. Quality improvement efforts aiming to improve interpersonal communication and provider attitudes, such as health provider trainings and mentorship, as well as leadership engagement may be promising avenues to enhance women’s experience of childbirth care in resource-constrained settings such as urban informal settlements in sub-Saharan Africa.

## Background

Sub-Saharan Africa is witnessing a rapid urbanization rate: 56% of the world’s population and almost 40% of its children are projected to live in African cities by 2050.^1^ Currently, 23% of the urban African population lives below the international poverty line, and 29% experience multidimensional poverty.^2^ Urban poor dwellers in informal settlements face precarious living conditions, characterized by overcrowding, congestion, inadequate hygiene and food security, air pollution, violence, and crime, among other health hazards. Moreover, high population density leads to overcrowded health facilities and limited capacity of human and physical resources to meet health demands.^3^ While it is expected that urban residence improves access to maternal, newborn and child health services through better infrastructure, challenging environments act as a bottleneck for urban poor women who are limited by geographic and financial access to sprawling informal facilities and sub-optimal quality services; ^3–5^ cities thus become a place where opportunity and adversity collide.

Recent data suggest that service contact with health services has increased in African capital cities, including institutional births reaching a median record of 95%.^3,6^ However, the content of care is variable, and maternal and newborn health outcomes remain disproportionately unfavorable among the urban poor.^7^ This sheds light on the gap between service contact and positive health outcomes,^3^ and the possible erosion of the once called “capital city advantage”.^8^ Similarly, systematic analyses suggest that the majority of deaths in low-and middle-income countries (LMICs) are due to poor quality of care,^9^ further highlighting the need for high quality services among marginalized populations.

Maternal and newborn quality of care measures have historically focused on assessing the provision of life-saving interventions, but these do not reflect women’s experiences of care. Yet, there is a growing body of evidence on neglectful, abusive and disrespectful treatment of women during childbirth.^10–16^ In Kenya, estimates suggest that about 20% of women leaving postnatal wards experienced some kind of mistreatment.^13^ In a qualitative study in Nairobi’s informal settlements, women reported verbal and physical abuse, poor rapport with providers, and feelings of neglect and abandonment during childbirth, indicating providers’ failures to meet professional standards.^11^ Similarly, observations of labor in Ghana, Guinea, Myanmar and Nigeria revealed that 42% of women experienced a form of physical or verbal abuse, stigma or discrimination around the time of delivery. Further evidence suggested it disproportionately affected women of low socio-economic status and adolescents girls. ^11,17–19^ Importantly, negative experiences of childbirth care can have multiplicative effects and significantly reduce subsequent utilization of health services,^20,21^ impacting health outcomes. This is of particular concern in sub-Saharan Africa, bearing the largest burden of maternal and newborn mortality globally.^22^

Despite this evidence, there has been a lack of consensus around the definitions and measures of childbirth care experience. In an effort to standardize and operationalize them, Afulani and colleagues developed and validated a person-centered maternity care (PCMC) scale in the context of LMICs in 2017, covering elements of dignity and respect, communication and autonomy, and supportive care during childbirth.^20^ PCMC refers to maternity care that is “respectful of and responsive to women’s and their families’ preferences, needs and values, and ensuring that their values guide all clinical decisions”.^23^ This definition employs a human rights-based approach to care and goes beyond mistreatment during childbirth, highlighting “holistic, responsive and dignified maternity care”,^20^ which is essential for continued care-seeking.

Yet, limited evidence exists on the experience of PCMC among marginalized urban communities. Our study objective was therefore to evaluate the experience of PCMC among women living in urban informal settlements in sub-Saharan African capital cities. Specifically, we aimed to 1) measure the levels of PCMC during childbirth and 2) identify structural, intermediary and health systems determinants associated with PCMC in Nairobi, Lusaka and Ouagadougou cities.

## Methods

### Study setting

This study was implemented between January and March 2023 in select urban informal settlements in Nairobi, Lusaka and Ouagadougou. These were locally defined geographic areas housing the urban poorest women in each city. Informal settlements are defined as areas in which residents lack tenure security, are cut off from basic services and infrastructure, and where housing does not comply with safety regulations, leading to hazardous conditions.^24^ In our study, while free maternity care policies exist in all three cities, it is important to recognize that each context is unique and the characteristics of populations living in urban informal settlements in Nairobi may be quite different from those in Lusaka or Ouagadougou.

We selected the following urban informal settlements based on existing collaborations and research activities: Korogocho and Viwandani in Nairobi, Chawama, Kanyama, George, Chipata, Chazanga, Chainda, Mtendere and Bauleni in Lusaka, and the East and West “non-loti” areas in Ouagadougou.

In 2019 approximately half of Nairobi’s 4.3 million inhabitants were informal settlement dwellers, yet these areas occupied only 5% of the city’s land.^25^ Korogocho is Nairobi’s fourth largest and most congested informal settlement with over 250 dwellings per hectare bordering the city’s main dumping site,^26^ and Viwandani is located in the industrial area.^27^ In Lusaka, close to 70% of the population live in informal settlements located in flood prone areas unauthorized for human housing, within industrial areas or the outskirts of the city.^28,29^ A previous study suggests these residents represent Lusaka’s 60% poorest households.^30^ Ouagadougou has a population of 2.4 million inhabitants and is divided into “loti” (formal) and “non-loti” (informal) areas. In 2022, 35% of Burkina Faso’s urban population lived in non-loti areas.^31^

### Study design

This cross-sectional study included a client exit survey of eligible women of reproductive age (15-49 years) who were discharged from delivery care in public and private health facilities identified as serving the select urban informal settlements in each city; health facility selection was based on previous mapping exercises and formative research. The survey was administered using distinct modalities: In Ouagadougou, we visited the client’s household within a week of facility discharge for an in-person survey. In Nairobi and Lusaka, we used two survey modalities: a phone-based survey within 1-2 weeks of discharge or an in-person client exit survey held in the facility in an area with maximum auditory and visual privacy. In Nairobi, the latter was conducted immediately upon facility discharge, whereas in Lusaka it was conducted upon completion of the postnatal care (PNC) visit within 1-2 weeks of delivery discharge.

The inclusion criteria were as follows: being an adult woman of reproductive age (18-49 years) or an emancipated minor (a married woman aged 15-17 years), living in the selected urban informal settlement (or its immediate neighborhood), and being discharged from childbirth care in a health facility primarily serving the urban informal settlement. Unmarried minors were excluded as they couldn’t provide informed consent; women who experienced a severe complication requiring referral or experienced a stillbirth or neonatal death were also excluded.

### Sample size and sampling

We calculated a sample size of 418 clients in each city to detect a mean PCMC score of 66.9% with a standard deviation of 13.6% (from previous studies in urban Kenya),^32^ with a margin of error of 3 percentage points (pp) and significance level (alpha) of 5%. In these urban settings, we assumed a non-response rate of 15% and a design effect of 4.5 to account for large clustering of women at the facility-level.

We selected a random sample of clients from all health facilities offering delivery care and serving our study sites. The allocation of clients was proportional to the expected delivery volume obtained from the facilities prior to the study start. Fieldworkers visited each facility daily to assess the eligibility of women using a screening form, and all eligible participants were recruited until the desired sample size was reached. In Nairobi and Lusaka where we implemented the dual survey modality, we first recruited women for the phone-based sample and then for the in-person survey which was administered at the health facility. In Ouagadougou, all eligible women were recruited in the facility upon discharge and followed-up for an in-person survey in their homes within a week.

### Measures

The client exit survey included questions on women’s demographic and socio-economic characteristics, antenatal care (ANC), care during labor and the immediate postpartum period, experience of PCMC, and overall satisfaction of care during labor and delivery. We used a validated PCMC scale which included 30 items across three core domains: dignity and respect, communication and autonomy, and supportive care. ^32,33^ The tool was adapted to each context and translated into local languages (Swahili in Nairobi, Bemba and Nyanja in Lusaka and Moore in Ouagadougou). We pre-tested the tool during the training and pilot study. The questionnaire was administered electronically using tablets and took approximately 20-30 minutes.

We generated the PCMC outcome following the methodology developed by Afulani et al.,^20,32^ whereby a score ranging from 0 to 3 was assigned to each of the 30 items (supplemental table 1). Responses were on a 4-point scale (no never, yes a few times, yes most of the time, yes all the time) and the highest numeric value of 3 was attributed to the response option reflecting optimal person-centered behavior. Values for all 30 items were summed, for a maximum score of 90 points for the full PCMC scale reflecting ideal person-centered care; a high score therefore reflects better PCMC. We also generated domain-specific scores for each of the three domain subscales: dignity and respect (6 items out of 30, for a total of 18 points), communication and autonomy (9 items for a total of 27 points), and supportive care (15 items for a total of 45 points). For ease of interpretation as a percentage and to allow comparison across domains,^32^ we rescaled the overall and domain scores to 100, though we maintained the unscaled scores in the regression models.

Following this study’s conceptual framework (supplemental figure 1), drawing elements from WHO’s Social Determinants of Health Framework and the Framework for Maternal and Newborn Care in Health Facilities,^34,35^ we identified structural, intermediary and health systems determinants of PCMC. Structural determinants reflect women’s socio-economic status and were measured using their education level and employment status in the last three months. We defined intermediary determinants as those depicting biological and behavioral factors, such as: women’s age, marital status, parity, experience of pregnancy complications, history of miscarriage/stillbirth, number of ANC visits during the last pregnancy, and place of ANC with respect to place of delivery. Lastly, we defined health systems determinants as the delivery facility type/level, facility managing authority, provider assistance during delivery, length of stay in facility for delivery, and we also included provision of care elements such as receipt of following before facility discharge: maternal PNC check, PNC counseling on danger signs, PNC counseling on family planning, PNC blood pressure measurement, newborn PNC check, and receipt of subsequent newborn PNC appointment.

### Statistical analysis

To identify factors associated with levels of PCMC, we explored bivariate associations and ran multilevel linear regression models, for each city, of the unscaled PCMC score (a continuous outcome out of 90 points) on women’s structural, intermediary and health systems determinants. Only complete cases were retained in models; missing values were <3%.

Since multiple women were recruited from a given health facility, and to account for facility-level clustering, we ran a two-level random intercept model with women as level 1 and facilities as level 2. Exploratory analyses for the linear regression included an assessment of normality of the unscaled PCMC scores by plotting a histogram of the PCMC scores as well as Q-Q plots of the residuals (supplemental figure 2).

We built the models sequentially for each city, following the order of our study conceptual framework (supplemental figure 1) to assess the effects of structural, intermediary, and health systems determinants. Model 1 tested associations between PCMC and structural determinants. In model 2, we added the intermediary determinants to model 1, and in the final model 3, we added the health systems determinants. We ran model 3 for the overall unscaled PCMC score outcome as well as for each unscaled PCMC domain score separately (dignity and respect, communication and autonomy, and supportive care).

Per Rabe-Hesketh and Skrondal,^36^ in the models below, *β_x_* through *β_z_* are the coefficients of interest that quantify the associations between each determinant and the PCMC score after adjusting for covariates. *ζ_j_* is the random intercept or level-2 residual, and represents the random deviation of facility *j*’s mean PCMC score from the overall mean PCMC score, accounting for between-facility variation, whereas ɛ*_ij_* is the woman-specific error component. We used variance inflation factors (VIF) to assess multicollinearity and correlation matrices to assess correlations between variables. In our final models, the VIF was of 1.32 in Nairobi, 1.37 in Lusaka and 1.42 in Ouagadougou, all of which were below the threshold of 5 to 10,^37^ indicating no multicollinearity.

In each city, for woman *i* and facility *j*

Model 1: structural determinants

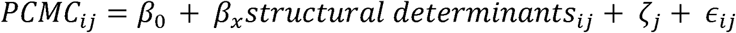

Model 2: structural and intermediary determinants

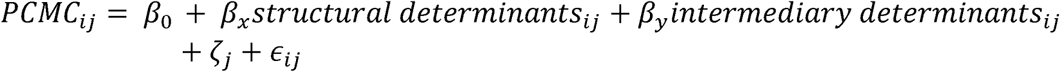

Model 3: structural, intermediary and health systems determinants

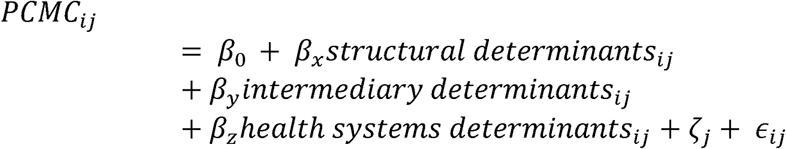

As a sensitivity analysis, we fit model 3 using a multilevel logistic regression: we defined PCMC as a dichotomous outcome using a cut-off score of 60 out of 90, reflecting the second most optimal response option to each of the 30 questions. The findings were consistent with those from our linear model (supplemental table 2).

### Ethical considerations

This study underwent ethical review and received approvals from in-country ethical review boards in each study site (AMREF approval P1309-2022and NACOSTI research license 686471 in Kenya, Comité d’éthique pour la Recherche en Santé approval 2022-10-216 in Burkina Faso, and University of Zambia Biomedical Research Ethics Committee approval 3215-2022 in Zambia), in addition to Johns Hopkins University IRB (approvals 22309 & 22304).

### Patient and public involvement

This study included women residents of urban informal settlements in Nairobi, Lusaka and Ouagadougou who were discharged from delivery care in a health facility. Prior to the implementation of the study, formative research was conducted with women residents of the study area who gave birth in the previous year. Women’s associations and representatives of informal settlement residents were involved in the interpretation of study results through dissemination meetings in each study site.

## Results

### Sample characteristics

#### Socio-economic and demographic characteristics

A total of 1,249 eligible women were included in the study: 412 in Nairobi, 436 in Lusaka and 401 in Ouagadougou. The response rate varied from 80.9% in Ouagadougou to 88.4% in Lusaka and 93.4% in Nairobi; reasons for non-response included refusal to participate and loss-to-follow-up. In Lusaka, all women were recruited from public facilities, compared to 76.5% and 65.8% in Nairobi and Ouagadougou, respectively. In Nairobi and Lusaka, 52.7% of women in our sample gave birth in a hospital, compared to 30.4% in Ouagadougou. Most women in Nairobi and Lusaka had secondary level education, whereas in Ouagadougou 42.1% had no formal education. Over 65% of women across cities were either unemployed or engaged in informal labor in the previous three months (table 1).

**Table 1.**
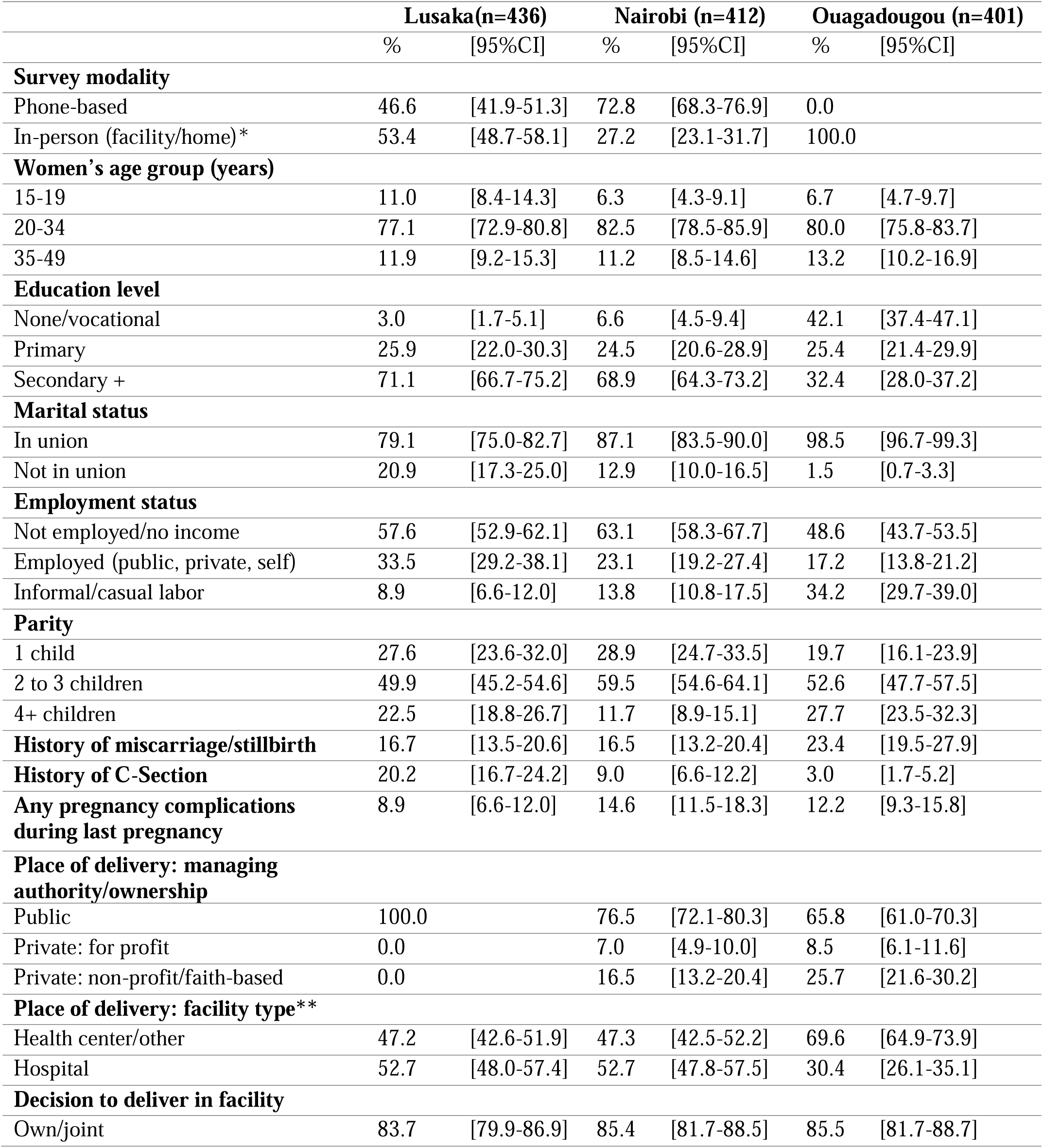
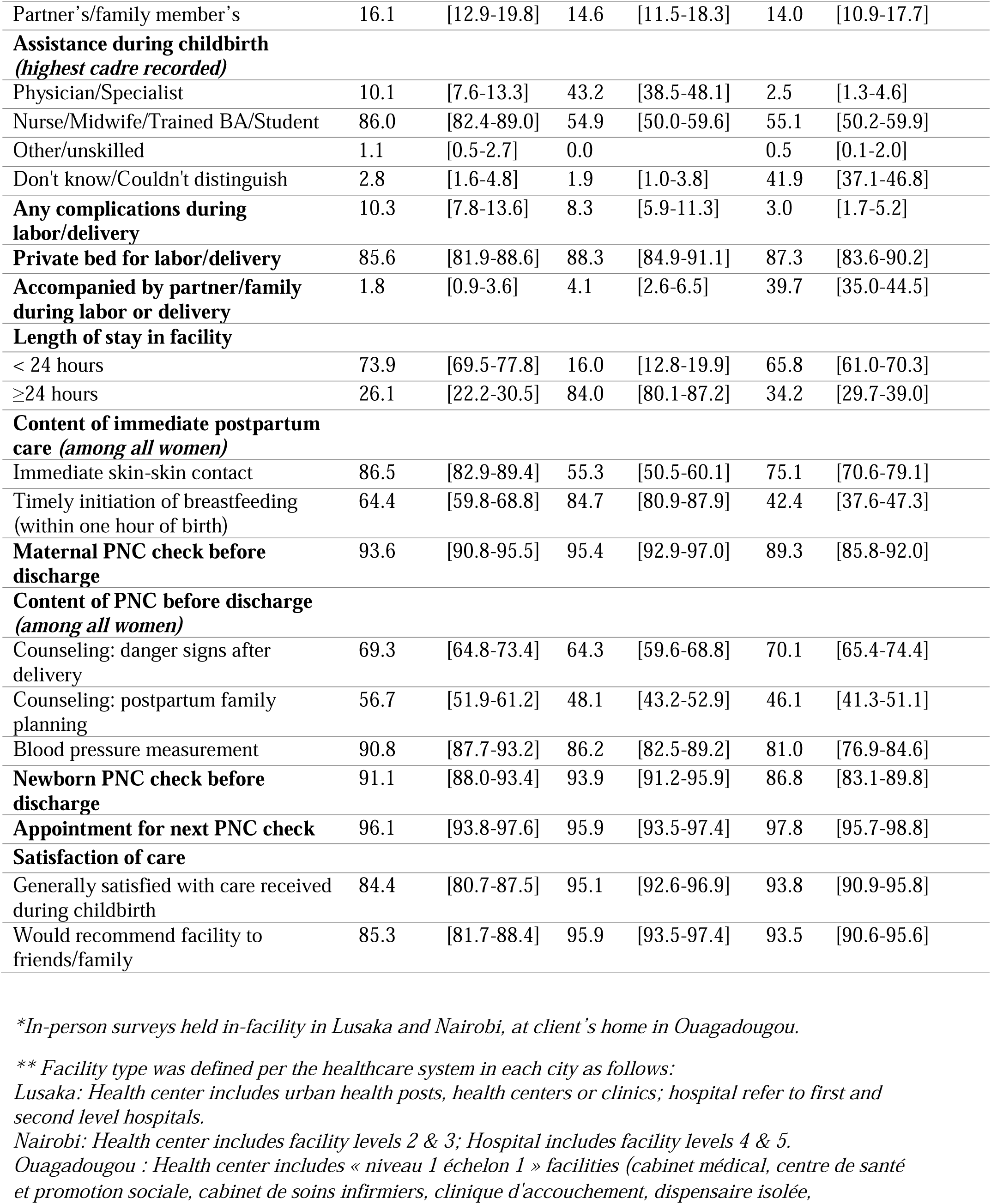
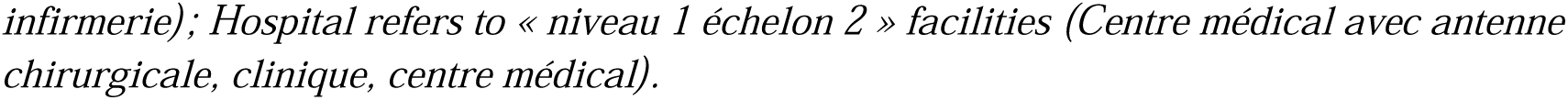
Women’s socio-economic, demographic characteristics and content of labor and delivery care by study site.

#### Antenatal care and childbirth care

In this urban context, more than 60% of women across sites received at least 4 ANC contacts during pregnancy, although fewer initiated ANC within the first trimester of gestation. Most women reported high content of ANC (supplemental table 3). In Nairobi, 43.2% of women were attended by a physician during childbirth, compared to 10.1% in Lusaka and 2.5% in Ouagadougou. Importantly, 41.9% of women in Ouagadougou were unable to distinguish the type of provider assisting them; this was below 3% in the other sites. While 39.7% of women in Ouagadougou reported having a birth companion, only 4.1% and 1.8% of women in Nairobi and Lusaka had one, respectively. Over 85% of women reported receiving a maternal or newborn PNC check before discharge; however, fewer women received counselling on pregnancy danger signs or postpartum family planning, across all cities (table 1).

### Level of person-centered maternity care

The overall mean PCMC score (rescaled) was 57.1% (95%CI 55.9, 58.4) in Lusaka, 69.5% (95%CI 68.4, 70.6) in Nairobi, and 73.8% (95%CI 72.6, 75.1) in Ouagadougou (figure 1). The score had an interquartile range of 48.8% to 65.6% in Lusaka, 61.1% to 77.8% in Nairobi, and 67.8% to 82.2% in Ouagadougou, where the median score of 76.6% was the highest (supplemental figure 3). No woman reported a perfect score of 100% in any city. Across sites, the mean dignity and respect domain scored highest, ranging from 57.1% (95%CI 55.9, 58.4) in Lusaka to 73.8% (95%CI 72.6, 75.1) in Ouagadougou, followed by supportive care. The lowest mean domain score was attributed to communication and autonomy, ranging from 47.6% (95%CI 45.6, 49.5) in Lusaka to 63.2% (95%CI 61.5, 64.9) in Nairobi (figure 1).

**Figure 1.**
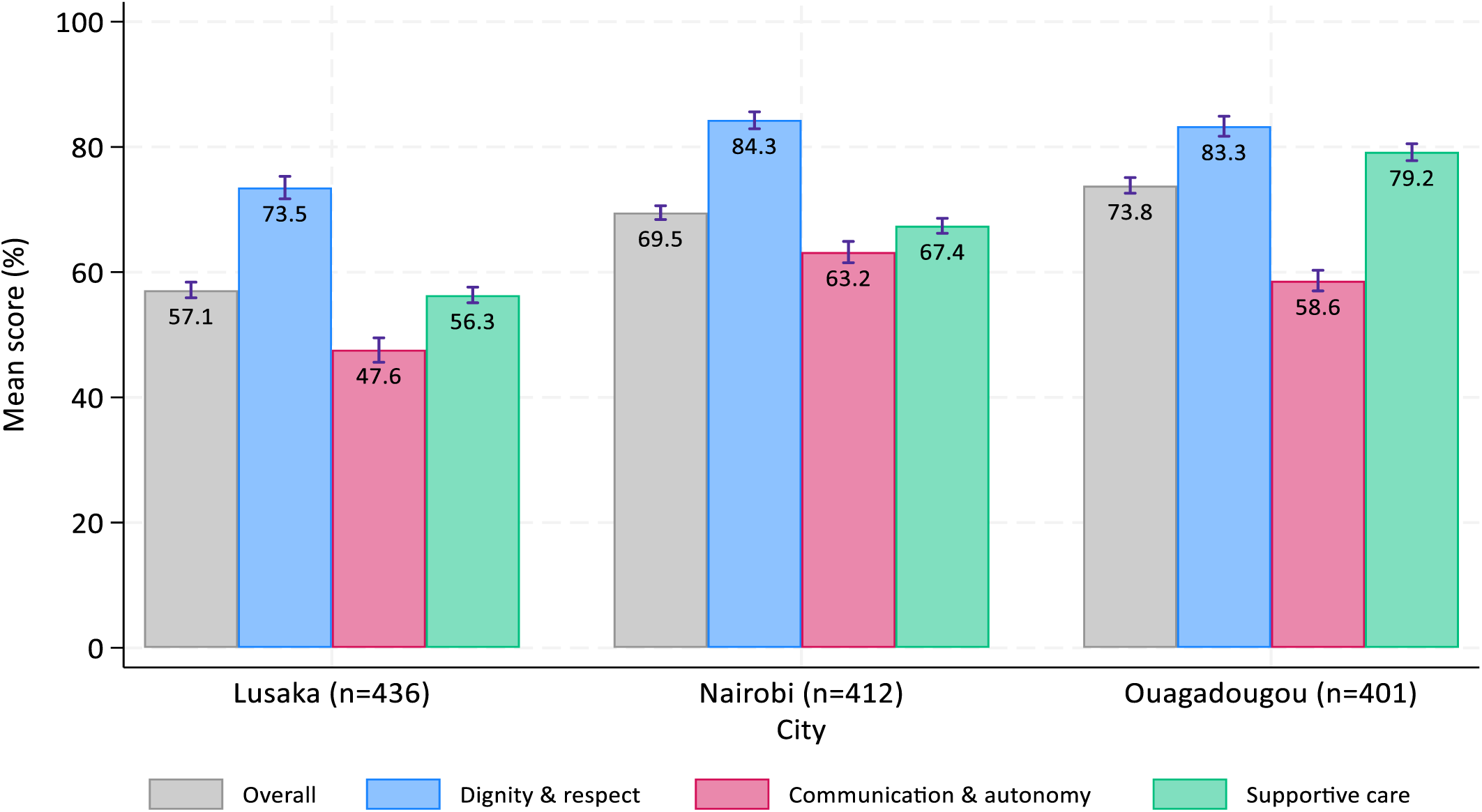
Overall and domain-specific PCMC scores (%) by study site.

Zooming in on specific items within PCMC domains, figure 2 and supplemental figures 4 A-B indicate that almost 23% and 6% of women in Lusaka experienced a form of verbal abuse (such as shouting, scolding, insulting or threatening), and physical abuse (treated roughly, pushed, beaten, slapped, pinched, etc.) during labor or delivery at least once, respectively. Similarly, over 70% of women in Lusaka and Ouagadougou, and 47% of women in Nairobi reported that providers never introduced themselves. Moreover, 57% and 68% of women in Lusaka and Nairobi respectively were never able to be in the position of their choice during delivery. While women generally felt supported during childbirth, 90% of women in Lusaka and 77% of those in Nairobi said they were never allowed to have a companion stay with them during labor; this was much lower at 19% in Ouagadougou.

**Figure 2:**
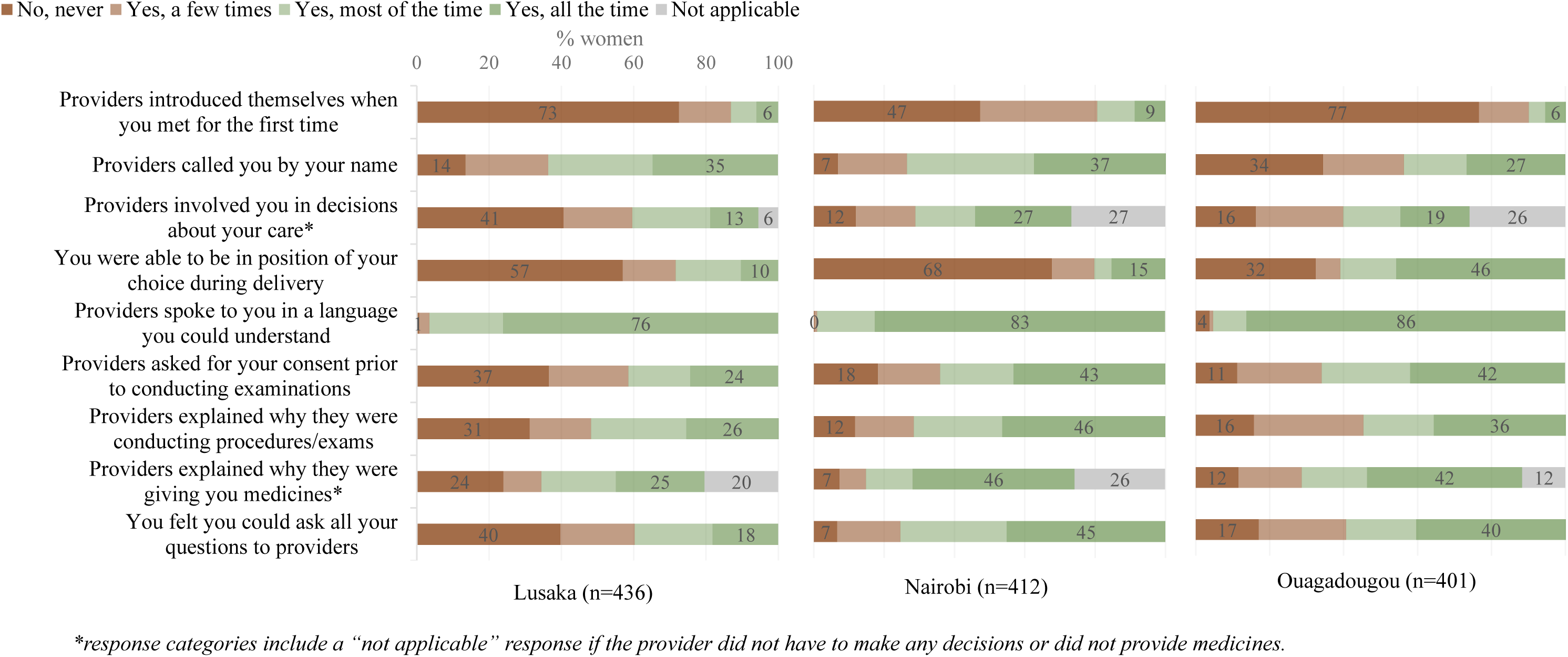
PCMC item responses (%) for communication and autonomy domain by study site.

The overall PCMC score varied by key socio-demographic characteristics: in Lusaka, formally employed women had a score of 59.3% compared to 55.9% among those unemployed. In Nairobi, women with secondary education reported a lower score than those with primary education. Similarly, women who delivered in a hospital reported a score of 64.3%, compared to 75.3% among those who attended health centers; the opposite was found in Ouagadougou. In Nairobi and Ouagadougou, women giving birth in public facilities reported a lower score than those who gave birth in private facilities (table 2).

**Table 2.**
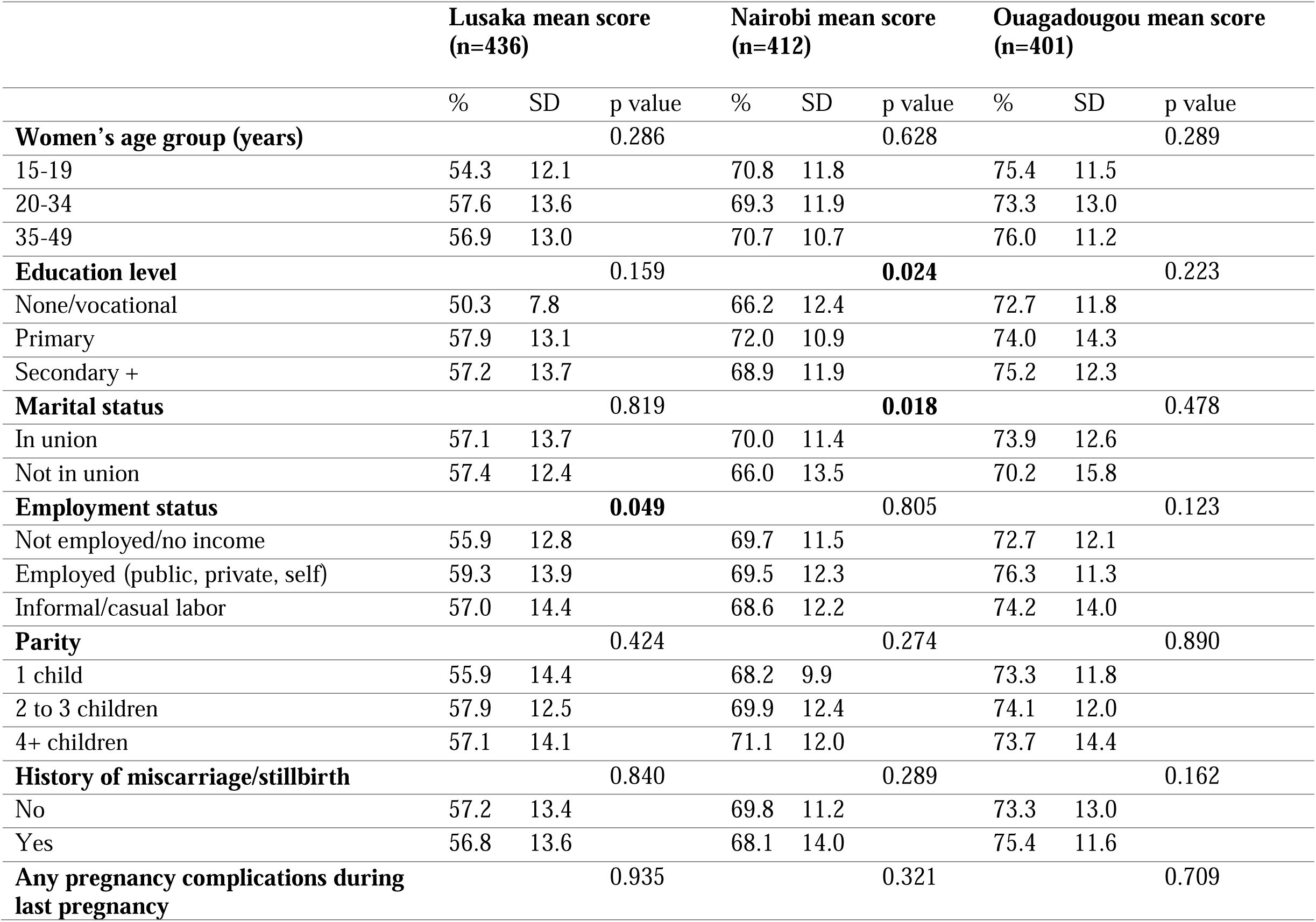
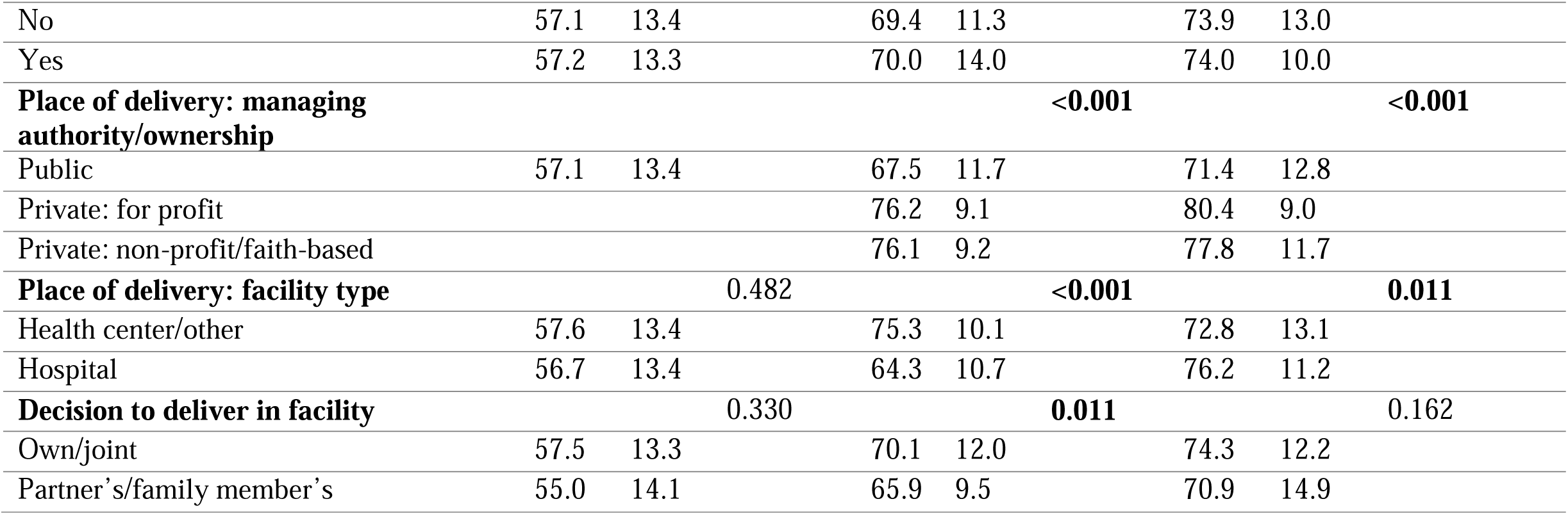
Overall PCMC scores (%) by women’s socio-economic and demographic characteristics.

### Determinants of person-centered maternity care

In Lusaka, model 3 including structural, intermediary and health systems determinants suggested that women who were attended by a physician compared to a nurse/midwife reported a statistically significant 4.5 pp (or 4.04 points out of 90) higher overall PCMC score (figure 3, supplemental table 5A), and they experienced better communication and autonomy by 9.5 pp (2.56 points out of 27) (supplemental table 6A). Women who received PNC counseling on danger signs reported a statistically significant 7.5pp (or 6.74 points out of 90) higher overall PCMC score (supplemental table 5A, figure 3), and this remained significant for each PCMC domain: increased dignity and respect score by 6.2 pp (1.12 points out of 18), better communication and autonomy by 9.6 pp (2.59 points out of 27), and better supportive care by 6.7 pp (3.02 points out of 45). Receipt of a newborn PNC check was associated with a higher overall reported PCMC score, and a statistically significantly better supportive care experience by 7.6 pp (3.41 points out of 45). Women with formal employment significantly experienced better communication compared to unemployed women (supplemental table 6A), although this association was not significant with overall PCMC in model 3 (figure 3, supplemental table 5A).

**Figure 3.**
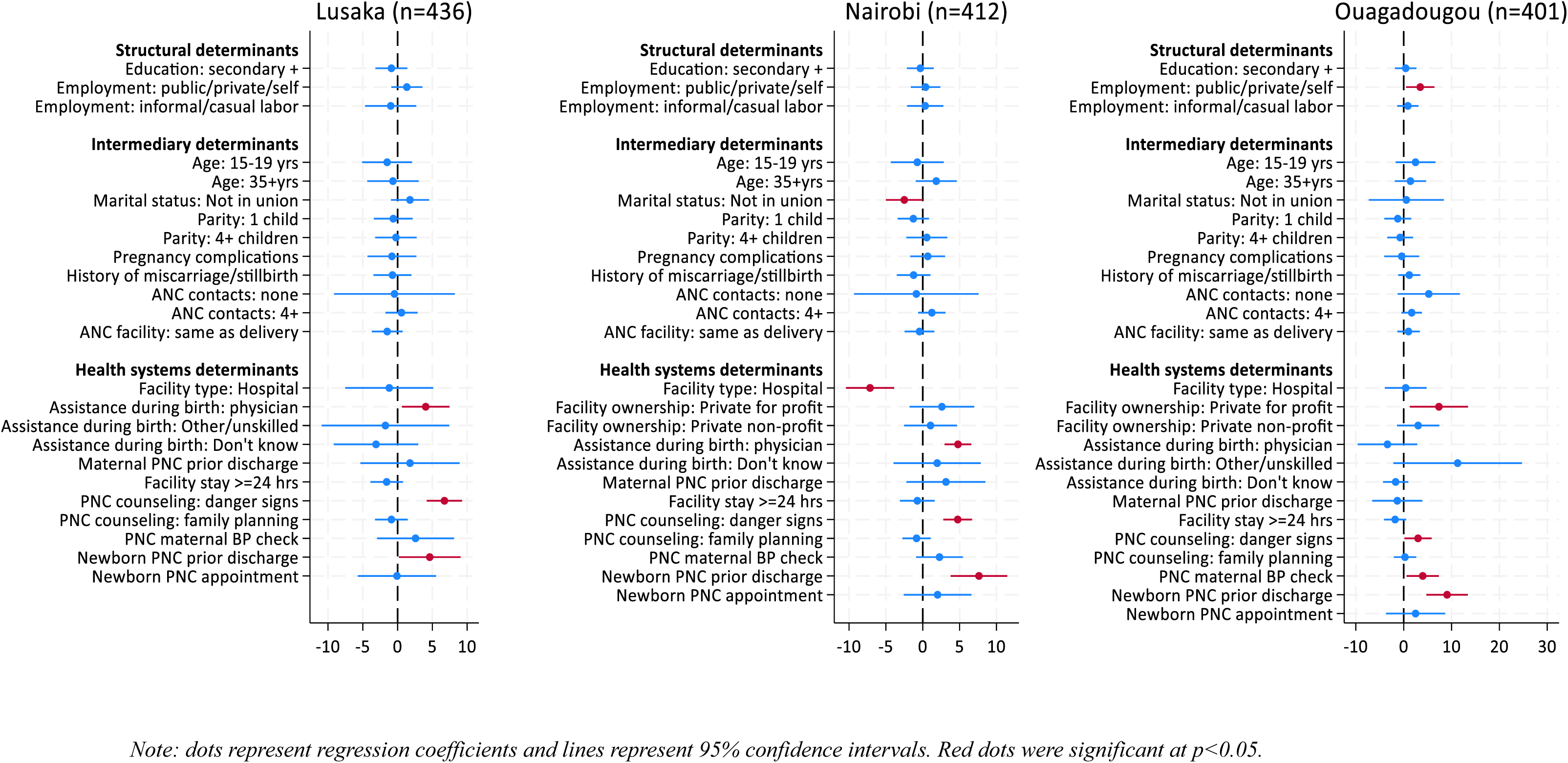
Structural, intermediary and health systems determinants of overall PCMC score (out of 90 points) by study site.

In Nairobi, the final model 3 suggested that married women experienced statistically significantly higher overall PCMC by 2.8 pp (2.5 points out of 90) (figure 3, supplemental table 5B); in particular, these women experienced better communication by 4.6 pp (1.24 points out of 27). Women aged 35-49 years reported better dignity and respect by 5.1 pp (0.91 points out of 18) compared to younger women aged 20-35 years (supplemental table 6B). In contrast, women who gave birth in a hospital compared to a lower-level facility reported a significantly poorer overall PCMC score by 7.9 pp; in particular, hospital births were associated with poorer communication and autonomy as well as supportive care by 7.0 pp and 10.7 pp, respectively.

That said, women who were attended by a physician, regardless of the place of delivery, had a 5.3 pp higher overall PCMC score (4.77 points out of 90), and this statistically significant positive effect persisted across all three PCMC domains. While there was no difference in overall PCMC score by facility managing authority, those who gave birth in private facilities reported a significantly higher dignity and respect score. As found in Lusaka, receipt of PNC counseling on danger signs and that of a newborn check were associated with higher overall reported PCMC, and this positive effect remained statistically significant for each PCMC domain (figure 3, supplemental tables 5B & 6B).

In Ouagadougou, the effect of employment was statistically significant in all models: formally employed women reported higher overall PCMC by 3.8 pp (3.45 points out of 90) compared to unemployed women in the final model (figure 3, supplemental table 5C). Further analyses revealed this was particularly associated with an improved communication and autonomy experience, by 7.2 pp (1.94 points out of 27) (supplemental table 6C). Women who gave birth in a private-for-profit facility reported an 8.2 pp higher overall PCMC than those in the public sector: specifically, they experienced better communication by 10.4 pp (2.8 points out of 27) and supportive care by 7.7 pp (3.46 points out of 45). Aligned with the Nairobi and Lusaka findings, women who received PNC counseling on danger signs reported a higher overall PCMC score, and specifically on the supportive care domain by 4.6 pp (2.05 points out of 45); those who received a blood pressure measurement experienced better communication by 9.5 pp (2.57 points out of 27). Finally, women who received a newborn PNC check reported a significant 10.1 pp higher overall PCMC score, as well as better scores for each PCMC domain (figure 3, supplemental tables 5C & 6C).

## Discussion

This study evaluates the levels and determinants of PCMC among 1,249 women living in urban informal settlements in the capital cities of Nairobi, Lusaka and Ouagadougou. Our analysis reveals inadequate levels of PCMC, with overall mean scores of 57.1% in Lusaka, 69.5% in Nairobi and 73.8% in Ouagadougou. Our estimates are consistent, though slightly higher than previously reported PCMC scores of 66.9% in urban Kenya,^32^ and 64.7% in similar informal settlements in Nairobi.^38^ Aligned with most studies in sub-Saharan Africa, we found poor patient-provider communication, with the majority of women reporting that providers did not explain the purpose of medical procedures, nor did they introduce themselves. ^32,38,39^ However, we found large variability in women’s ability to have a birth companion in Ouagadougou compared to Lusaka and Nairobi, and this is often influenced by the facility’s policy.

Additionally, we found that 15% and 2% of women in Nairobi experienced verbal abuse and physical abuse at least once respectively, in line with other quantitative findings in urban Kenya.^32,38^ This was even more prevalent in our Lusaka study, with 23% and 6% of women reporting verbal and physical abuse, respectively. Evidence from Kenya suggests that mistreatment during childbirth is driven by factors occurring at multiple levels: individual, family, community and health systems.^12,40^ These may include normalized behaviors in low-resource settings, imbalanced provider-patient power dynamics, bias, as well as poor facility infrastructure, health worker stress/burnout, demotivation, and heavy workload in crowded wards.^11,12,40^ Further research is needed to understand health providers’ barriers and enablers in offering PCMC in low-resource settings. Our study indicates that despite existing frameworks and policies promoting respectful maternity care and person-centered practices in Kenya, such as the Quality Model for Health and the National Reproductive Health Policy,^41,42^ the agenda remains unfinished, especially among marginalized urban populations.

Our analysis identifying structural, intermediary and health systems factors associated with PCMC revealed important differences across sites pointing to the unique context in each one, but there were also some similarities. In general, we found that select health systems factors had strong associations with PCMC in the urban informal settlement context: birth assistance by a physician compared to a nurse/midwife significantly improved levels of PCMC in Nairobi and Lusaka, particularly in areas of patient-provider communication. Our findings are consistent with Stierman et al. reporting a signification correlation between receipt of a postpartum check before discharge and PCMC in Ethiopia.^39^ Provision of care, as measured in our study by self-reported receipt of PNC components before discharge, was significantly associated with better PCMC across all cities. The PCMC scale covers elements of care from women’s first interaction with health providers through their time in the postnatal ward, and immediate postpartum/postnatal care occurs downstream from other components of care captured. Therefore, this result could reflect unmeasured upstream characteristics associated with higher PCMC. It is also possible that positive interactions during PNC may influence women’s reporting of their overall experience of care, as these events are temporally closest to the time of discharge and survey participation; however, more research is needed to unpack these findings.

Furthermore, we found that women who gave birth in a hospital in Nairobi reported lower overall PCMC and poorer communication and supportive care than those from lower-level facilities. Our supplementary analyses indicated that a larger proportion of women in hospitals compared to health centers in Nairobi reported overcrowding, and fewer felt safe and trusted the providers (supplemental table 7). In line with our findings, studies in Kenya and Ethiopia revealed that women delivering in hospitals were more likely to report mistreatment during childbirth, including physical abuse and neglect.^17,32^ While higher level facilities are generally better equipped with physical and human resources, they may be less conducive towards offering PCMC given large patient volumes, understaffing and high provider stress/burnout.^17^ Health system redesign strategies aiming to shift deliveries to higher-level facilities ^43^ ought to ensure that providers are supported to offer PCMC, as negative experiences during childbirth can have detrimental effects on women’s continuity in service use.^44^ In Ouagadougou, our findings of higher reported PCMC in private for-profit facilities complement those from a qualitative study identifying the need to promote physical and social environments in public health facilities in Burkina Faso to enhance patients’ experiences.^45^

Our study uncovered few structural and intermediary determinants of PCMC among urban informal settlement residents, even at the bivariate level, possibly explained by low variability in women’s socio-economic characteristics in this setting. That said, previous findings have pointed to poorer experiences of care and a higher likelihood of abuse among poorer women.^17,46^ There have been conflicting results on the effect of education on PCMC: while we did not find a significant effect, other studies in Kenya including one in informal settlements in Nairobi found higher PCMC reporting among literate and wealthier women.^32,38^ Similarly, in Ethiopia, younger age (under 20 years), lower wealth, not achieving 4 ANC visits, and experiencing complications during pregnancy were associated with lower PCMC. Moreover, the authors found wide variation in PCMC even within communities, suggesting that provider-patient interactions may be unique to each individual, rather than to a community.^39^

Despite growing evidence of inadequate PCMC in sub-Saharan Africa, particularly among urban informal settlement dwellers as evidenced in our study, a dearth of interventions exist to address this issue, and most are limited to provider training on respectful maternity care.^47–49^ Implementation research in Kenya led to key recommendations to promote respectful maternity care: a participatory approach engaging stakeholders, community sensitization, and respective maternity care workshops complementing routine health provider trainings, including counseling and supportive management structures for providers.^50^ One such intervention, conducted between 2011 and 2014 in Kenya, involved a three-tiered approach at policy, facility and community-levels, and led to reductions in the frequency of disrespect and abuse during childbirth of up to 20%.^51^ Similarly, a theory and evidence-based intervention by Afulani et al. in Kenya and Ghana focused on addressing two key drivers of poor PCMC: provider stress and implicit bias. It involved five integrated components: provider training, peer support, mentorship, embedded champions and leadership engagement.^40,52^ The recent evaluation of its pilot study in Migori county, Kenya, indicated significant declines in providers’ perceived stress and burnout following the intervention, and improvements in their well-being and stress management competences. Such integrated interventions have the potential to create an enabling environment to improve women’s experience of PCMC.^53^

Our study has important strengths: in addition to allowing multi-country comparisons, we focused on marginalized urban populations living in informal settlements who bear the largest mortality burden in cities and are too often left behind. Our findings build on the scientific literature in Kenya and generate actionable evidence in Zambia and Burkina Faso where the PCMC literature is sparse. There are also limitations worth mentioning: PCMC measurement relies on women’s self-report and is prone to recall bias. Evidence suggests that the accuracy of women’s recall on intrapartum care declines over time^54–56^; to address this, we minimized the recall period to 1-2 weeks following facility discharge. Despite its validation in LMICs, in the absence of a gold standard measure the PCMC score may underestimate disrespectful maternity care. Specific components of the PCMC scale are inherently subjective, and poor provider behavior may be normalized in low-resource settings;^39^ this is also true for perceptions of facility crowdedness and cleanliness. Women surveyed in the facility may encounter courtesy reporting bias.^14,57,58^ We explored this by comparing phone-based and in-person survey results in Nairobi and Lusaka; these results are described elsewhere. Our study did not include populations living in wealthier neighborhoods, and further comparative studies would be beneficial.

In conclusion, our study indicates that urban informal settlement dwellers in Nairobi, Lusaka and Ouagadougou experienced inadequate PCMC, particularly in areas of patient-provider communication and women’s autonomy. Most women reported that providers did not explain reasons for procedures nor sought consent prior to conducting examinations. While the majority of women felt supported and respected during childbirth, our study uncovered actionable gaps, such as ensuring women are covered up during examinations in the labor room, enabling them to have a birth companion and to deliver in their position of choice. Health systems factors such as facility type and managing authority, provider assisting at birth, and provision of postnatal care prior to discharge were significantly associated with PCMC in this resource-constrained setting. More research is needed to understand health providers’ barriers in offering PCMC and the structures enabling PCMC in this context. Quality improvement efforts and evidence-based interventions integrating provider training, peer support and leadership engagement ^52,53^ may be promising avenues to enhance women’s experience of childbirth care in informal settlements, and to ensure continuity in care seeking.

## Data Availability

All data produced in the present study are available upon reasonable request to the authors.

## Acknowledgements

We are immensely grateful to all the women who agreed to participate in our study and share their experiences of maternity care, as well as to their families for their support. We thank and acknowledge the hard work of study team members in Nairobi, Lusaka and Ouagadougou, in particular data collectors and field supervisors. We also thank facility staff and in-charges for their time. We recognize all team members and Countdown to 2030 colleagues who contributed towards the study design, implementation, analysis and dissemination. In addition, SS Jiwani is thankful to Dr. Melinda Munos and Dr. Elizabeth Stierman for their support and guidance through her Thesis Advisory Committee.

## Contributions

SS Jiwani was responsible for the overall content as guarantor. SS Jiwani, A Amouzou and C Faye conceptualized the study, with inputs from T Boerma. SS Jiwani developed the first version of study tools, with review from K Cisse, C Jacobs, M Mutua. The study was implemented by SS Jiwani, K Cisse, C Jacobs, M Mutua, A Njeri, G Adero, M Musukuma, D Ngosam and F Sissoko. SS Jiwani lead the statistical analysis and developed the first draft of the manuscript with inputs from A Amouzou. S Kouanda provided technical advice and review; A Abajobir supported dissemination of findings. All co-authors reviewed and contributed to the final version of this manuscript.

## Data availability

The datasets generated and/or analyzed during the current study can be made available from the corresponding author on reasonable request.

