## Supplementary materials for "Levels and Determinants of Person-Centered Maternity Care Among Women Living in Urban Informal Settlements: Evidence from Client Exit Surveys in Nairobi, Lusaka and Ouagadougou"

### SUPPLEMENTAL MATERIALS

Supplemental table 1. PCMC scale questions, response categories and scoring

| Question | Response categories | PCMC scoring (out of 90) |
|---|---|---|
| **DIGNITY & RESPECT** | | |
| Did the doctors, nurses, or other staff at the facility treat you with respect? | No, never | 0 |
| | Yes, a few times | 1 |
| | Yes, most of the time | 2 |
| | Yes, all the time | 3 |
| Did the doctors, nurses, or other staff at the facility treat you in a friendly manner? | No, never | 0 |
| | Yes, a few times | 1 |
| | Yes, most of the time | 2 |
| | Yes, all the time | 3 |
| Did you feel that they shouted at you, scolded, insulted, threatened, or talked to you rudely? | No, never | 3 |
| | Yes, once | 2 |
| | Yes, a few times | 1 |
| | Yes, many times | 0 |
| | Refused to respond | 3 |
| Did you feel like you were treated roughly like pushed, beaten, slapped, pinched, physically restrained, or gagged? | No, never | 3 |
| | Yes, once | 2 |
| | Yes, a few times | 1 |
| | Yes, many times | 0 |
| | Refused to respond | 3 |
| During examinations in the labor room, were you covered up with a cloth or blanket, or screened with a curtain so that you did not feel exposed? | No, never | 0 |
| | Yes, a few times | 1 |
| | Yes, most of the time | 2 |
| | Yes, all the time | 3 |
| Do you feel like your health information was or will be kept confidential at this facility? | No, never | 0 |
| | Yes, a few times | 1 |
| | Yes, most of the time | 2 |
| | Yes, all the time | 3 |
| **COMMUNICATION & AUTONOMY** | | |
| During your time in the health facility did the doctors, nurses, or other health-care providers introduce themselves to you when they first came to see you? | No, none of them | 0 |
| | Yes, a few of them | 1 |
| | Yes, most of them | 2 |
| | Yes, all of them | 3 |

| | | |
|---|---|---|
| Did the doctors, nurses, or other health-care providers call you by your name? | No, never | 0 |
| | Yes, a few times | 1 |
| | Yes, most of the time | 2 |
| | Yes, all the time | 3 |
| Did you feel like the doctors, nurses or other staff at the facility involved you in decisions about your care? | No, never | 0 |
| | Yes, a few times | 1 |
| | Yes, most of the time | 2 |
| | Yes, all the time | 3 |
| | Did not have to make any decisions | 3 |
| During the delivery, do you feel like you were able to be in the position of your choice? | No, never | 0 |
| | Yes, for a short time | 1 |
| | Yes, most of the time | 2 |
| | Yes, all the time | 3 |
| Did the doctors, nurses, or other staff at the facility speak to you in a language you could understand? | No, never | 0 |
| | Yes, a few times | 1 |
| | Yes, most of the time | 2 |
| | Yes, all the time | 3 |
| Did the doctors, nurses, or other staff at the facility ask your permission or consent before doing procedures on you? | No, never | 0 |
| | Yes, a few times | 1 |
| | Yes, most of the time | 2 |
| | Yes, all the time | 3 |
| Did the doctors, nurses, or other staff at the facility explain to you why they were doing examinations or procedures on you? | No, never | 0 |
| | Yes, a few times | 1 |
| | Yes, most of the time | 2 |
| | Yes, all the time | 3 |
| Did the doctors, nurses, or other staff at the facility explain to you why they were giving you any medicine? | No, never | 0 |
| | Yes, a few times | 1 |
| | Yes, most of the time | 2 |
| | Yes, all the time | 3 |
| | Did not get any medicine | 3 |
| Did you feel you could ask the doctors, nurses, or other staff at the facility any questions you had? | No, never | 0 |
| | Yes, a few times | 1 |
| | Yes, most of the time | 2 |
| | Yes, all the time | 3 |
| **SUPPORTIVE CARE** | | |
| How did you feel about the amount of time you waited to receive care? Would you say it was: | Very short | 3 |
| | Somewhat short | 2 |
| | Somewhat long | 1 |
| | Very long | 0 |
| Did the doctors and nurses at the facility show concern for your feelings about your delivery? | No, never | 0 |
| | Yes, a few times | 1 |
| | Yes, most of the time | 2 |

| | | |
|---|---|---|
| | Yes, all the time | 3 |
| Did the doctors, nurses, or other staff at the facility try to understand your anxieties? | No, never | 0 |
| | Yes, a few times | 1 |
| | Yes, most of the time | 2 |
| | Yes, all the time | 3 |
| | Did not have any anxiety | 3 |
| When you needed help, did you feel the doctors, nurses, or other staff at the facility paid attention? | No, never | 0 |
| | Yes, a few times | 1 |
| | Yes, most of the time | 2 |
| | Yes, all the time | 3 |
| Do you feel the doctors or nurses did everything they could to help control your pain? | No, never | 0 |
| | Yes, a few times | 1 |
| | Yes, most of the time | 2 |
| | Yes, all the time | 3 |
| Were you allowed to have someone you wanted (outside of staff at the facility, such as family or friends) to stay with you during labor? | No, never | 0 |
| | Yes, a few times | 1 |
| | Yes, most of the time | 2 |
| | Yes, all the time | 3 |
| | I did not want anyone to stay with me | 3 |
| Were you allowed to have someone you wanted to stay with you during delivery? | No, never | 0 |
| | Yes, a few times | 1 |
| | Yes, most of the time | 2 |
| | Yes, all the time | 3 |
| | I did not want anyone to stay with me | 3 |
| Did you feel the doctors, nurses, or other staff at the facility took good care of you, at the best of their ability? | No, never | 0 |
| | Yes, a few times | 1 |
| | Yes, most of the time | 2 |
| | Yes, all the time | 3 |
| Did you feel you could completely trust the doctors, nurses, or other staff at the facility with regards to your care? | No, never | 0 |
| | Yes, a few times | 1 |
| | Yes, most of the time | 2 |
| | Yes, all the time | 3 |
| Do you think there were enough health staff in the facility to care for you? | No, never | 0 |
| | Yes, a few times | 1 |
| | Yes, most of the time | 2 |
| | Yes, all the time | 3 |
| Thinking about the labor and postnatal wards, did you feel the health facility was crowded? | No, never | 3 |
| | Yes, a few times | 2 |
| | Yes, most of the time | 1 |
| | Yes, all the time | 0 |
| | Very dirty | 0 |

| | | |
|---|---|---|
| Thinking about the wards, washrooms, and the general environment of the health facility, would you say the facility was very clean, clean, dirty, or very dirty? | Dirty | 1 |
| | Clean | 2 |
| | Very clean | 3 |
| Was there running water in the facility? | No, never | 0 |
| | Yes, a few times | 1 |
| | Yes, most of the time | 2 |
| | Yes, all the time | 3 |
| Was there electricity in the facility? | No, never | 0 |
| | Yes, a few times | 1 |
| | Yes, most of the time | 2 |
| | Yes, all the time | 3 |
| In general, did you feel safe in the health facility? | No, never | 0 |
| | Yes, a few times | 1 |
| | Yes, most of the time | 2 |
| | Yes, all the time | 3 |

Supplemental figure 1. Study conceptual framework

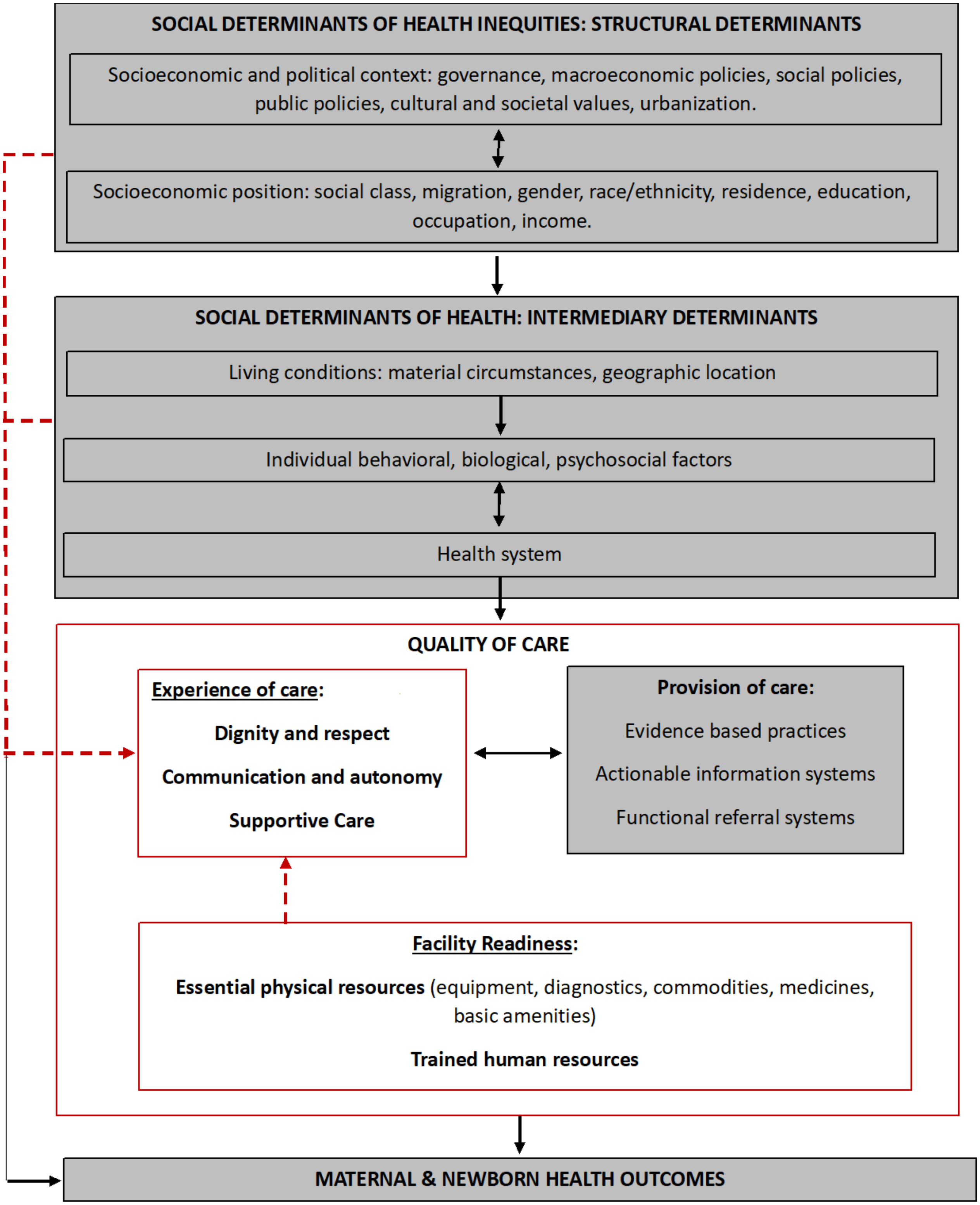

Supplemental figure 2. Histogram and Q-Q plot of unscaled PCMC score (out of 90) by study site

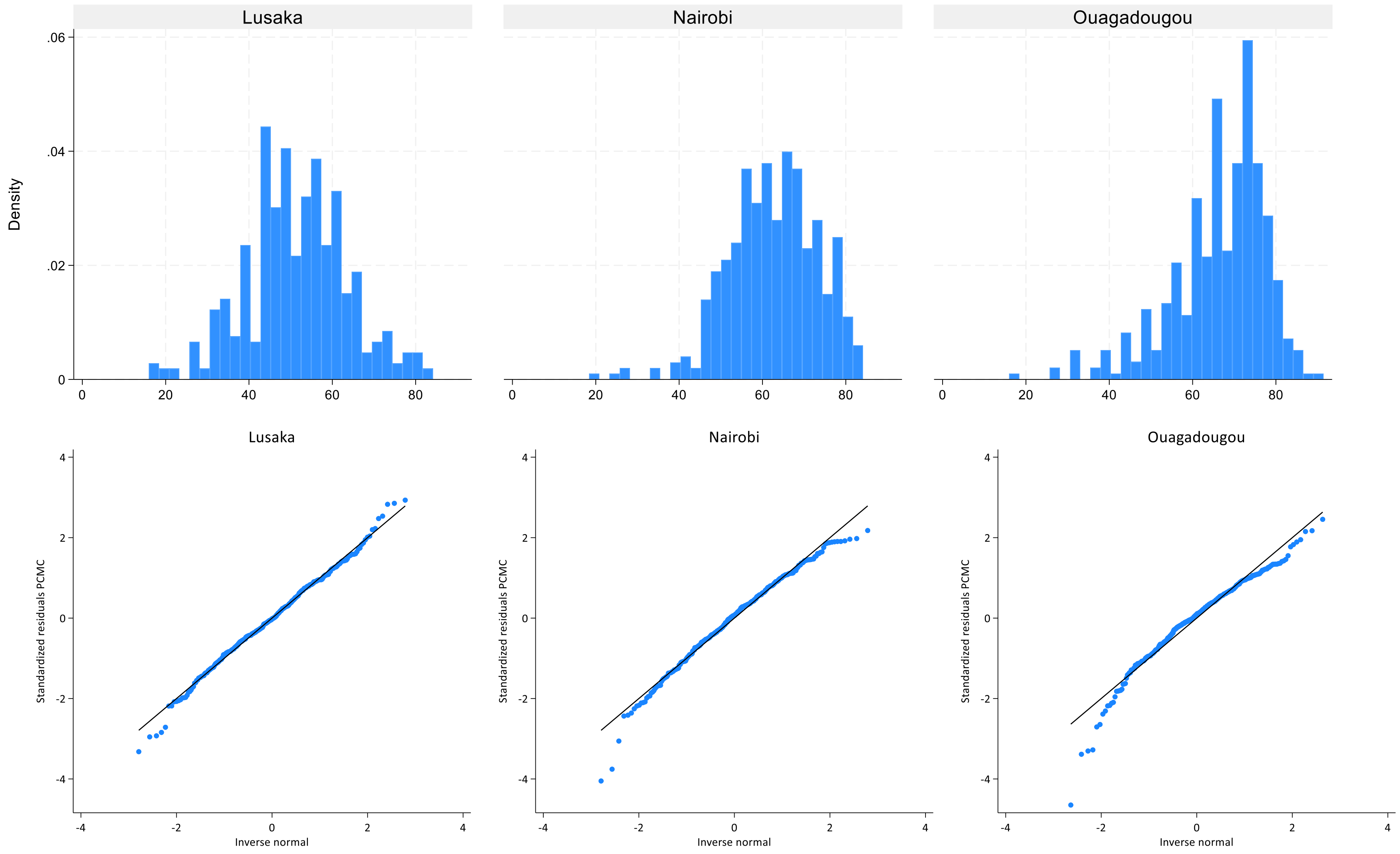

Supplemental table 2. Sensitivity analysis: PCMC logistic regression by study site.

| | LUSAKA Odds Ratio | NAIROBI Odds Ratio | OUAGADOUGOU Odds Ratio |
|---|---|---|---|
| *STRUCTURAL DETERMINANTS* | | | |
| **Education** (ref: primary or less) | | | |
| Secondary or more | 1.12 | 0.99 | 0.88 |
| | [0.60,2.10] | [0.58,1.69] | [0.37,2.09] |
| **Employment** (ref: unemployed) | | | |
| Employed (public, private, self) | 0.91 | 1.13 | **5.08*** |
| | [0.50,1.63] | [0.61,2.08] | **[1.47,17.49]** |
| Informal/casual labor | 1.68 | 1.00 | 1.10 |
| | [0.62,4.55] | [0.48,2.08] | [0.48,2.48] |
| *INTERMEDIARY DETERMINANTS* | | | |
| **Age** (ref: 20-35 yrs) | | | |
| 15-19 yrs | 0.44 | 0.93 | 1.16 |
| | [0.16,1.25] | [0.33,2.65] | [0.24,5.67] |
| 35-49 yrs | 1.21 | 1.66 | 1.22 |
| | [0.46,3.19] | [0.71,3.87] | [0.34,4.34] |
| **Marital status** (ref: in union) | | | |
| Not in union | **2.27*** | 0.86 | 0.74 |
| | **[1.09,4.72]** | [0.42,1.76] | [0.06,9.46] |
| **Parity** (ref: 2-3 children) | | | |
| 1 | 1.15 | 0.64 | 0.90 |
| | [0.56,2.38] | [0.34,1.19] | [0.32,2.55] |
| 4+ | 0.86 | 0.67 | 0.67 |
| | [0.38,1.94] | [0.30,1.49] | [0.25,1.83] |
| **Pregnancy complications** (ref: No) | | | |
| Yes | 0.63 | 1.55 | 1.25 |
| | [0.21,1.89] | [0.73,3.30] | [0.31,5.00] |
| **Miscarriage/Stillbirth history** (ref: No) | | | |
| Yes | 0.65 | 0.73 | 1.65 |
| | [0.30,1.39] | [0.37,1.45] | [0.69,3.98] |
| **Number of ANC contacts** (ref: 1-3) | | | |
| 0 | 1.21 | 0.13 | 4.20 |
| | [0.16,9.05] | [0.01,1.60] | [0.25,70.69] |
| 4+ | 0.91 | 1.39 | 1.19 |
| | [0.49,1.69] | [0.80,2.43] | [0.54,2.63] |
| **Place of ANC** (ref: Different facility/home/no ANC) | | | |
| Same facility as place of delivery | 0.87 | 0.82 | 0.92 |
| | [0.49,1.56] | [0.45,1.50] | [0.39,2.17] |
| *HEALTH SYSTEMS DETERMINANTS* | | | |
| **Delivery facility type** (ref: Health center) | | | |
| Hospital | 1.20 | **0.24**** | 1.30 |
| | [0.25,5.83] | **[0.12,0.49]** | [0.26,6.52] |
| **Delivery facility managing authority/ownership** (ref: Public) | | | |
| Private for profit | - | 1.99 | 6.50 |

| | | | |
|---|---|---|---|
| | | [0.55,7.12] | [0.50,84.75] |
| Private non-profit/faith-based | - | 1.75 | 2.01 |
| | | [0.64,4.78] | [0.38,10.49] |
| **Assistance during delivery** (ref: midwife /nurse /TBA) | | | |
| Physician/specialist | 1.67 | **2.62**$^{**}$ | 0.27 |
| | [0.70,3.99] | **[1.55,4.43]** | [0.03,2.77] |
| Other/unskilled | 1.27 | - | - |
| | [0.15,10.49] | | |
| Don't Know/Couldn't distinguish | 0.29 | 2.15 | 0.48 |
| | [0.03,2.90] | [0.42,10.98] | [0.18,1.27] |
| **Maternal PNC before discharge** (ref: No) | | | |
| Yes | - | 0.68 | 0.41 |
| | | [0.12,3.83] | [0.07,2.49] |
| **Length of facility stay** (ref: <24h) | | | |
| ≥24h | 0.54 | 0.98 | **0.32**$^{*}$ |
| | [0.28,1.02] | [0.47,2.07] | **[0.13,0.77]** |
| **PNC counseling: danger signs** (ref: No) | | | |
| Yes | **2.44**$^{*}$ | **3.81**$^{**}$ | 2.57 |
| | **[1.19,4.98]** | **[2.17,6.69]** | [0.94,7.02] |
| **PNC counselling: family planning** (ref: No) | | | |
| Yes | 0.81 | 0.86 | 0.88 |
| | [0.44,1.49] | [0.50,1.48] | [0.34,2.27] |
| **PNC: BP check** (ref: No) | | | |
| Yes | 4.09 | 0.96 | 1.46 |
| | [0.43,38.48] | [0.29,3.11] | [0.38,5.59] |
| **PNC: newborn check** (ref: No) | | | |
| Yes | - | **8.33**$^{**}$ | **8.22**$^{**}$ |
| | | **[2.35,29.62]** | **[1.77,38.29]** |
| **PNC: newborn appointment** (ref: No) | | | |
| Yes | 0.59 | 0.71 | 4.01 |
| | [0.10,3.53] | [0.19,2.69] | [0.64,24.92] |
| Observations (n) | 377 | 405 | 369 |

95% confidence intervals in brackets
$^{*}$ $p < 0.05$, $^{**}$ $p < 0.01$

Supplemental table 3. Care seeking and content of antenatal care during pregnancy by study site.

| | **Lusaka (n=436)** | | **Nairobi (n=412)** | | **Ouagadougou (n=401)** | |
|---|---|---|---|---|---|---|
| | % | [95%CI] | % | [95%CI] | % | [95%CI] |
| **Place of ANC** | | | | | | |
| Home/no ANC | 1.8 | [0.9-3.6] | 1.0 | [0.4-2.6] | 2.5 | [1.3-4.6] |
| Same health facility as place of delivery | 62.8 | [58.2-67.3] | 33.2 | [28.9-38.0] | 71.1 | [66.4-75.3] |
| Other health facility | 35.3 | [31.0-39.9] | 65.8 | [61.0-70.2] | 26.4 | [22.3-31.0] |
| **Place of ANC: facility type** | | | | | | |
| Home/no ANC | 1.8 | [0.9-3.6] | 1.0 | [0.4-2.6] | 2.5 | [1.3-4.6] |
| Hospital | 51.8 | [47.1-56.5] | 24.3 | [20.4-28.7] | 26.9 | [22.8-31.5] |
| Health center/other | 46.3 | [41.6-51.0] | 74.8 | [70.3-78.7] | 70.6 | [65.9-74.8] |
| **Number of ANC contacts** | | | | | | |
| No ANC | 1.4 | [0.6-3.0] | 1.0 | [0.4-2.6] | 2.5 | [1.3-4.6] |
| 1-3 contacts | 30.0 | [25.9-34.5] | 32.5 | [28.2-37.2] | 27.2 | [23.0-31.8] |
| 4-7 contacts | 61.9 | [57.3-66.4] | 62.9 | [58.1-67.4] | 63.8 | [59.0-68.4] |
| 8+ contacts | 6.7 | [4.7-9.4] | 3.4 | [2.0-5.7] | 1.2 | [0.5-3.0] |
| **Timely ANC initiation (first trimester)** | 22.3 | [18.6-26.5] | 23.8 | [19.9-28.2] | 31.7 | [27.3-36.5] |
| **ANC content** | | | | | | |
| Blood pressure | 99.1 | [97.5-99.7] | 99.8 | [98.3-100.0] | 97.7 | [95.6-98.8] |
| Blood sample | 91.4 | [88.3-93.7] | 98.8 | [97.1-99.5] | 92.6 | [89.5-94.8] |
| Urine sample | 72.1 | [67.6-76.1] | 98.5 | [96.8-99.3] | 64.5 | [59.6-69.1] |
| HIV test | 97.9 | [96.0-98.9] | 99.5 | [98.1-99.9] | 74.7 | [70.1-78.8] |
| Counseling: pregnancy danger signs | 88.6 | [85.2-91.3] | 88.5 | [85.0-91.2] | 69.6 | [64.8-73.9] |
| Counseling: nutrition | 94.0 | [91.3-95.9] | 86.5 | [82.8-89.5] | 52.2 | [47.2-57.1] |
| Any IFA Supplementation | 94.4 | [91.8-96.2] | 76.7 | [72.3-80.6] | 98.2 | [96.3-99.1] |
| Any IPTp (SP/Fansidar) | 95.1 | [92.6-96.8] | 31.9 | [27.5-36.6] | 97.7 | [95.6-98.8] |
| Tetanus toxoid vaccine | | | | | 83.9 | [79.9-87.2] |
| **ANC content score* (median, IQR)** | 100 | [87.5-100] | 87.5 | [75.0- 87.5] | 77.8 | [66.7-88.9] |

**ANC score defined as the number of ANC content interventions received, out of the number of interventions collected, expressed as a percentage.*

Supplemental figure 3: Distribution of overall PCMC scores (%) by study site

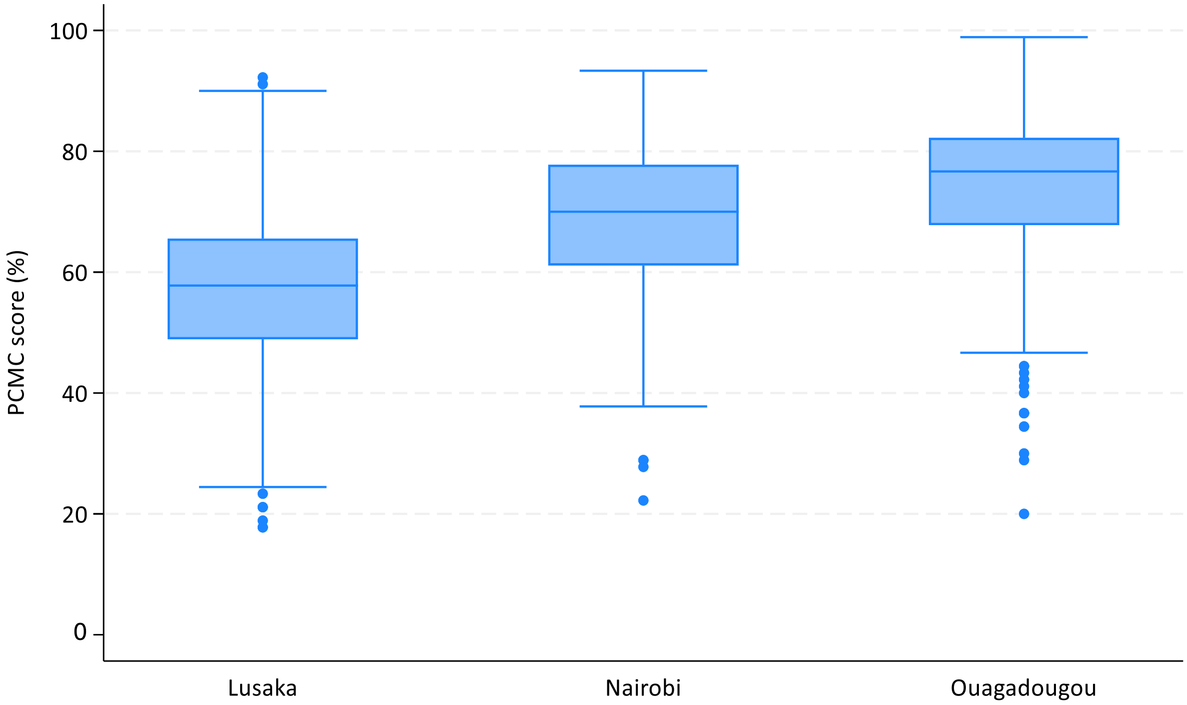

Supplemental figures 4 A-B. PCMC item responses (%) for dignity and respect (A), and supportive care (B) domains by study site.

A. Dignity and respect

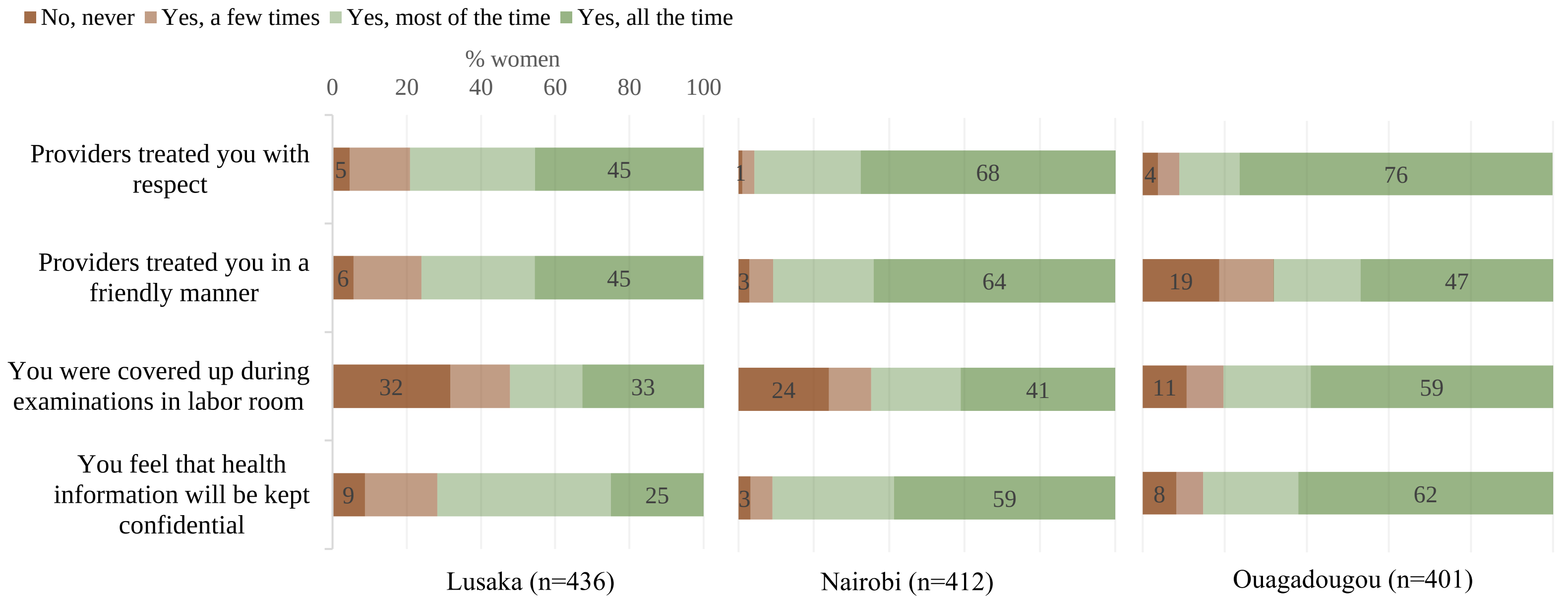


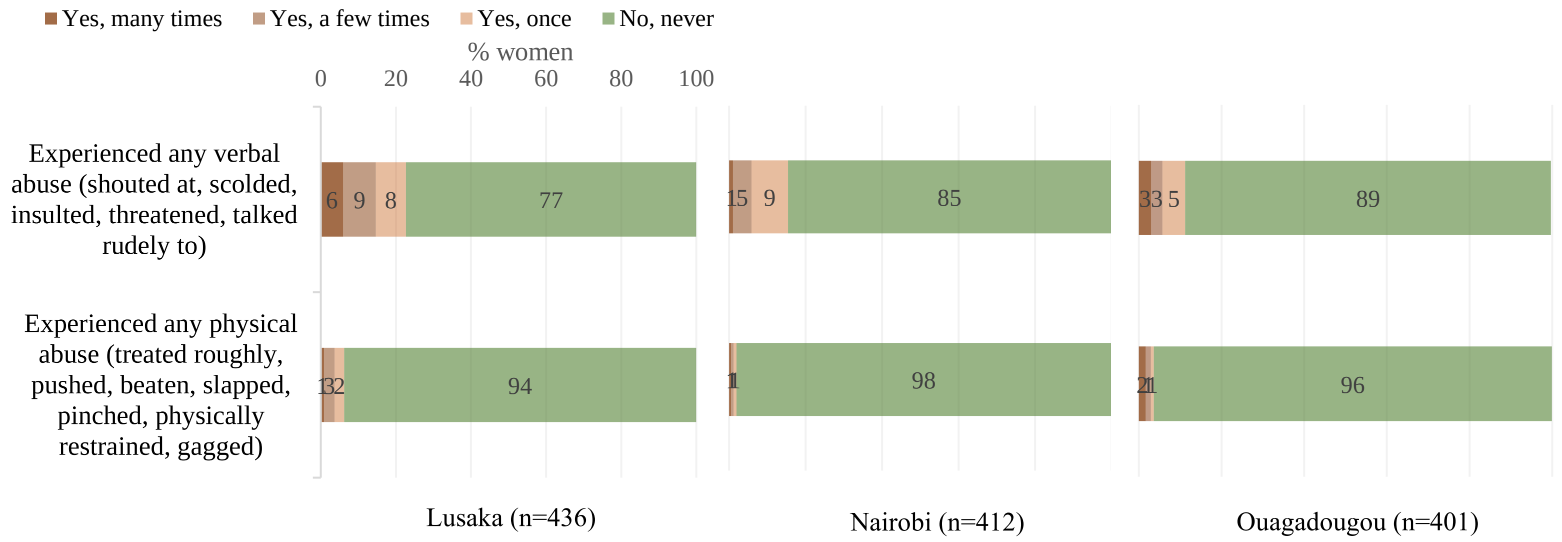

B. Supportive care

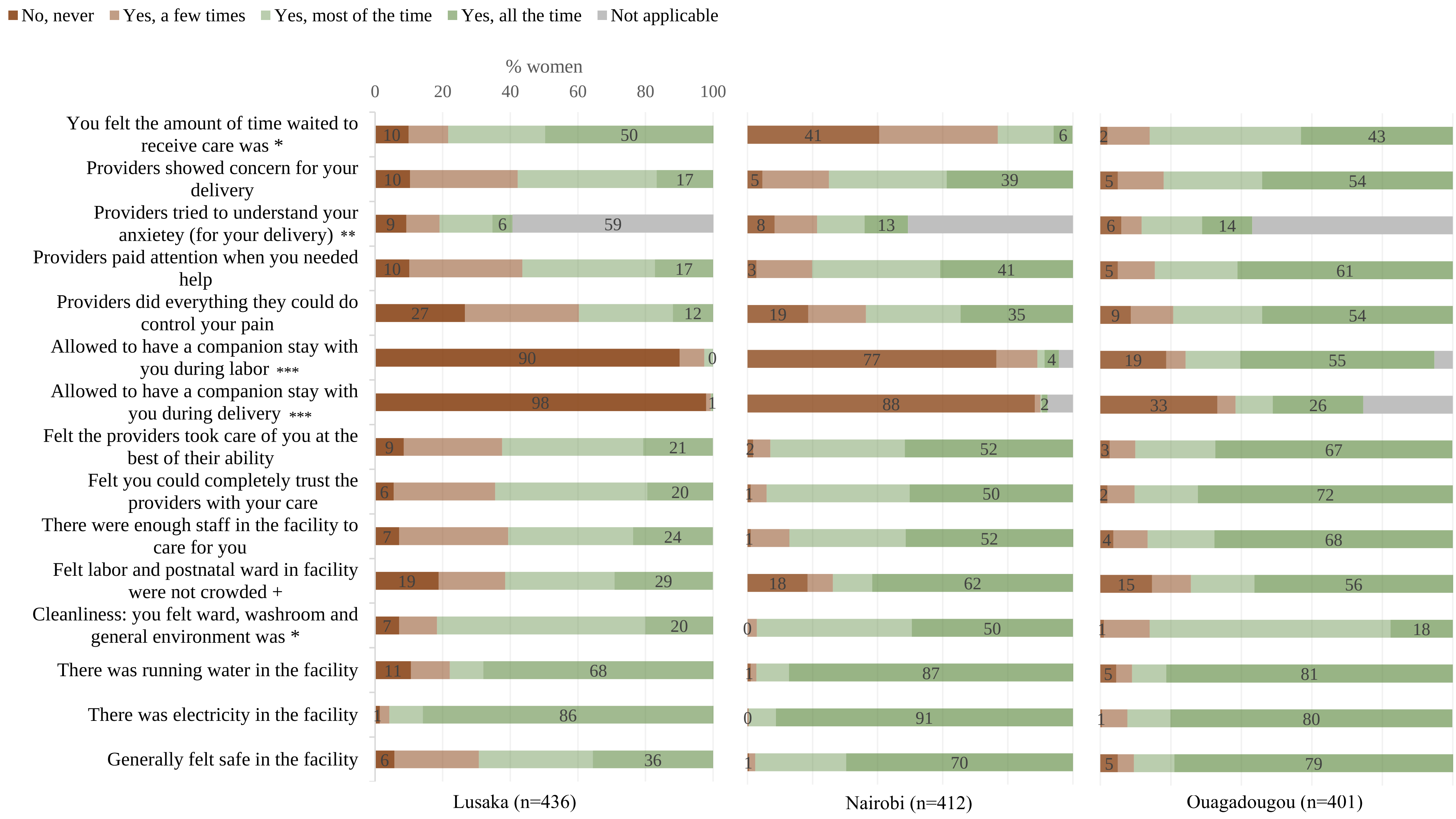

**response categories differ:*
*amount of time waited: very long (dark red), somewhat long, somewhat short, very short (dark green);*
*cleanliness: very dirty (dark red), dirty, clean, very clean (dark green).*
*** Not applicable response: "I was not feeling anxious".*
**** Not applicable response: "I did not want anyone to stay with me".*
*+ crowdedness: crowded all the time (dark red), most of the time, a few times, never crowded (dark green).*

Supplemental tables 5 A-C. Structural, intermediary and health systems determinants of overall PCMC by study site.

A. Lusaka: Structural, intermediary, health systems determinants of PCMC (unscaled score, out of 90)

| | Bivariate | Model 1 | Model 2 | Model 3 |
|---|---|---|---|---|
| | *STRUCTURAL DETERMINANTS* | | | |
| **Education** (ref: primary or less) | | | | |
| Secondary or more | 0.30<br>[-2.01,2.61] | 0.17<br>[-2.13,2.48] | 0.31<br>[-2.10,2.71] | -0.89<br>[-3.18,1.40] |
| **Employment** (ref: unemployed) | | | | |
| Employed: public, private, self | **2.66***<br>**[0.40,4.91]** | **2.65***<br>**[0.39,4.91]** | **2.49***<br>**[0.18,4.80]** | 1.33<br>[-0.89,3.55] |
| Informal/casual labor | 0.25<br>[-3.58,4.07] | 0.27<br>[-3.57,4.10] | -0.04<br>[-3.91,3.84] | -0.98<br>[-4.65,2.69] |
| | *INTERMEDIARY DETERMINANTS* | | | |
| **Age** (ref: 20-35 yrs) | | | | |
| 15-19 yrs | -2.23<br>[-5.56,1.10] | | -0.87<br>[-4.70,2.97] | -1.50<br>[-5.09,2.09] |
| 35-49 yrs | -0.75<br>[-4.01,2.50] | | -0.24<br>[-4.10,3.62] | -0.67<br>[-4.36,3.03] |
| **Marital status** (ref: in union) | | | | |
| Not in union | 0.24<br>[-2.35,2.83] | | 1.32<br>[-1.58,4.22] | 1.79<br>[-0.96,4.53] |
| **Parity** (ref: 2-3 children) | | | | |
| 1 | -1.76<br>[-4.22,0.71] | | -1.97<br>[-4.87,0.92] | -0.61<br>[-3.41,2.18] |
| 4+ | -0.93<br>[-3.57,1.72] | | -0.80<br>[-3.93,2.33] | -0.24<br>[-3.21,2.73] |
| **Pregnancy complications** (ref: No) | | | | |
| Yes | 0.67<br>[-2.98,4.32] | | 0.33<br>[-3.32,3.97] | -0.81<br>[-4.32,2.71] |
| **Miscarriage/Stillbirth history** (ref: No) | | | | |
| Yes | -1.41<br>[-4.20,1.39] | | -1.82<br>[-4.68,1.04] | -0.73<br>[-3.46,2.00] |
| **Number of ANC contacts** (ref: 1-3) | | | | |
| 0 | 0.63<br>[-8.54,9.81] | | -0.33<br>[-9.61,8.94] | -0.44<br>[-9.12,8.24] |
| 4+ | 0.39<br>[-1.93,2.71] | | 0.31<br>[-2.09,2.72] | 0.57<br>[-1.73,2.87] |
| **Place of ANC** (ref: Different facility/home/no ANC) | | | | |
| Same facility as place of delivery | -0.56<br>[-2.78,1.66] | | -0.53<br>[-2.78,1.73] | -1.50<br>[-3.71,0.71] |
| | *HEALTH SYSTEMS DETERMINANTS* | | | |
| **Delivery facility type** (ref: Health | | | | |

| | | | | |
|---|---|---|---|---|
| center) | | | | |
| Hospital | -0.92 | | | -1.20 |
| | [-8.00,6.17] | | | [-7.52,5.13] |
| **Assistance during delivery** (ref: midwife/nurse/TBA) | | | | |
| Physician/specialist | 2.43 | | | **4.04**$^{*}$ |
| | [-1.06,5.92] | | | **[0.62,7.46]** |
| Other/unskilled | -1.62 | | | -1.75 |
| | [-11.45,8.21] | | | [-10.91,7.42] |
| Don't Know/ Could not distinguish | -4.28<br>[-10.64,2.08] | | | -3.10<br>[-9.16,2.97] |
| **Maternal PNC before discharge** (ref: No) | | | | |
| Yes | **10.73**$^{**}$ | | | 1.79 |
| | **[6.11,15.35]** | | | [-5.33,8.91] |
| **Length of facility stay** (ref: <24h) | | | | |
| ≥24h | 0.08 | | | -1.57 |
| | [-2.31,2.47] | | | [-3.91,0.77] |
| **PNC counseling: danger signs** (ref: No) | | | | |
| Yes | **7.81**$^{**}$ | | | **6.74**$^{**}$ |
| | **[5.63,9.98]** | | | **[4.20,9.28]** |
| **PNC counselling: family planning** (ref: No) | | | | |
| Yes | **3.12**$^{**}$ | | | -0.88 |
| | **[0.99,5.26]** | | | [-3.22,1.45] |
| **PNC: BP check** (ref: No) | | | | |
| Yes | **8.11**$^{**}$ | | | 2.59 |
| | **[4.50,11.72]** | | | [-2.95,8.13] |
| **PNC: newborn check** (ref: No) | | | | |
| Yes | **9.46**$^{**}$ | | | **4.63**$^{*}$ |
| | **[5.51,13.42]** | | | **[0.20,9.06]** |
| **PNC: newborn appointment** (ref: No) | | | | |
| Yes | 3.77 | | | -0.09 |
| | [-2.08,9.61] | | | [-5.71,5.54] |
| Observations (n) | | 436 | 434 | 419 |
| AIC | | 3358.3 | 3358.6 | 3200.3 |

95% confidence intervals in brackets
$^{*}$ $p < 0.05$, $^{**}$ $p < 0.01$

B. Nairobi: Structural, intermediary, health systems determinants of PCMC (unscaled score, out of 90)

| | Bivariate | Model 1 | Model 2 | Model 3 |
|---|---|---|---|---|
| *STRUCTURAL DETERMINANTS* | | | | |
| **Education** (ref: primary or less) | | | | |
| Secondary or more | -1.04 [-2.97,0.88] | -1.08 [-3.01,0.86] | -0.86 [-2.84,1.12] | -0.33 [-2.14,1.49] |
| **Employment** (ref: unemployed) | | | | |
| Employed: public, private, self | 0.22 [-1.93,2.37] | 0.27 [-1.88,2.42] | 0.07 [-2.09,2.24] | 0.39 [-1.61,2.38] |
| Informal/casual labor | -0.23 [-2.88,2.41] | -0.33 [-2.97,2.32] | -0.11 [-2.76,2.54] | 0.33 [-2.13,2.80] |
| *INTERMEDIARY DETERMINANTS* | | | | |
| **Age** (ref: 20-35 yrs) | | | | |
| 15-19 yrs | -0.15 [-3.82,3.52] | | -0.59 [-4.55,3.37] | -0.75 [-4.34,2.85] |
| 35-49 yrs | 1.96 [-0.86,4.78] | | 1.53 [-1.47,4.53] | 1.83 [-0.95,4.61] |
| **Marital status** (ref: in union) | | | | |
| Not in union | **-2.89*** **[-5.54,-0.25]** | | -2.56 [-5.29,0.18] | **-2.50*** **[-4.99,-0.01]** |
| **Parity** (ref: 2-3 children) | | | | |
| 1 | -1.08 [-3.09,0.92] | | -0.68 [-2.97,1.60] | -1.28 [-3.40,0.84] |
| 4+ | 0.25 [-2.64,3.13] | | -0.07 [-3.04,2.90] | 0.55 [-2.22,3.33] |
| **Pregnancy complications** (ref: No) | | | | |
| Yes | 0.17 [-2.36,2.69] | | 0.00 [-2.54,2.55] | 0.67 [-1.69,3.04] |
| **Miscarriage/Stillbirth history** (ref: No) | | | | |
| Yes | -1.00 [-3.39,1.39] | | -1.47 [-3.91,0.97] | -1.24 [-3.52,1.04] |
| **Number of ANC contacts** (ref: 1-3) | | | | |
| 0 | 1.30 [-7.78,10.37] | | 1.28 [-7.91,10.48] | -0.87 [-9.31,7.57] |
| 4+ | 0.39 [-1.53,2.30] | | 0.77 [-1.24,2.79] | 1.23 [-0.61,3.08] |
| **Place of ANC** (ref: Different facility/home/no ANC) | | | | |
| Same facility as place of delivery | 0.56 [-1.65,2.76] | | 0.39 [-1.80,2.59] | -0.44 [-2.45,1.56] |
| *HEALTH SYSTEMS DETERMINANTS* | | | | |
| **Delivery facility type** (ref: Health center) | | | | |
| Hospital | **-9.48**** | | | **-7.15**** |

| | | | | |
|---|---|---|---|---|
| | **[-13.11,-5.85]** | | | **[-10.42,-3.89]** |
| **Delivery facility managing authority/ownership** (ref: Public) | | | | |
| Private for profit | 5.39 | | | 2.59 |
| | [-1.39,12.16] | | | [-1.78,6.97] |
| Private non-profit/faith-based | 5.36 | | | 1.06 |
| | [-0.56,11.28] | | | [-2.53,4.64] |
| **Assistance during delivery** (ref: midwife/nurse/TBA) | | | | |
| Physician/specialist | **4.39**$^{**}$ | | | **4.77**$^{**}$ |
| | **[2.44,6.33]** | | | **[2.96,6.59]** |
| Don't Know/Could not distinguish | 1.71 | | | 1.96 |
| | [-4.61,8.02] | | | [-3.97,7.88] |
| **Maternal PNC before discharge** (ref: No) | | | | |
| Yes | **7.90**$^{**}$ | | | 3.14 |
| | **[3.49,12.31]** | | | [-2.21,8.50] |
| **Length of facility stay** (ref: <24h) | | | | |
| ≥24h | -0.69 | | | -0.74 |
| | [-3.23,1.86] | | | [-3.08,1.61] |
| **PNC counseling: danger signs** (ref: No) | | | | |
| Yes | **5.50**$^{**}$ | | | **4.74**$^{**}$ |
| | **[3.63,7.37]** | | | **[2.80,6.68]** |
| **PNC counselling: family planning** (ref: No) | | | | |
| Yes | **2.01**$^{*}$ | | | -0.85 |
| | **[0.03,3.98]** | | | [-2.76,1.07] |
| **PNC: BP check** (ref: No) | | | | |
| Yes | **4.07**$^{**}$ | | | 2.27 |
| | **[1.30,6.83]** | | | [-0.90,5.44] |
| **PNC: newborn check** (ref: No) | | | | |
| Yes | **9.31**$^{**}$ | | | **7.63**$^{**}$ |
| | **[5.54,13.08]** | | | **[3.80,11.45]** |
| **PNC: newborn appointment** (ref: No) | | | | |
| Yes | 4.28 | | | 2.01 |
| | [-0.34,8.90] | | | [-2.58,6.61] |
| Observations (n) | | 412 | 410 | 405 |
| AIC | | 3024.0 | 3017.6 | 2909.9 |

95% confidence intervals in brackets
$^{*}$ $p < 0.05$, $^{**}$ $p < 0.01$

C. Ouagadougou: Structural, intermediary, health systems determinants of PCMC (unscaled score, out of 90)

| | Bivariate | Model 1 | Model 2 | Model 3 |
|---|---|---|---|---|
| | *STRUCTURAL DETERMINANTS* | | | |
| **Education** (ref: primary or less) | | | | |
| Secondary or more | 0.69 | 0.15 | 0.35 | 0.41 |
| | [-1.49,2.87] | [-2.06,2.37] | [-2.14,2.83] | [-1.82,2.65] |
| **Employment** (ref: unemployed) | | | | |
| Employed (public, private, self) | **4.20**$^{**}$ | **4.17**$^{**}$ | **4.59**$^{**}$ | **3.45**$^{*}$ |
| | **[1.27,7.13]** | **[1.19,7.14]** | **[1.33,7.86]** | **[0.49,6.41]** |
| Informal/casual labor | 0.77 | 0.79 | 1.01 | 0.87 |
| | [-1.54,3.08] | [-1.54,3.12] | [-1.44,3.46] | [-1.34,3.07] |
| | *INTERMEDIARY DETERMINANTS* | | | |
| **Age** (ref: 20-35 yrs) | | | | |
| 15-19 yrs | 1.22 | | 2.77 | 2.47 |
| | [-2.88,5.32] | | [-1.78,7.32] | [-1.68,6.62] |
| 35-49 yrs | 2.23 | | 3.20 | 1.41 |
| | [-0.77,5.23] | | [-0.42,6.81] | [-1.87,4.68] |
| **Marital status** (ref: in union) | | | | |
| Not in union | -3.12 | | -0.63 | 0.54 |
| | [-11.59,5.36] | | [-9.22,7.97] | [-7.30,8.38] |
| **Parity** (ref: 2-3 children) | | | | |
| 1 | -1.35 | | -1.59 | -1.25 |
| | [-4.05,1.35] | | [-4.71,1.52] | [-4.05,1.56] |
| 4+ | -0.14 | | -1.31 | -0.75 |
| | [-2.55,2.26] | | [-4.30,1.67] | [-3.43,1.94] |
| **Pregnancy complications** (ref: No) | | | | |
| Yes | 0.50 | | 0.39 | -0.43 |
| | [-3.40,4.40] | | [-3.57,4.35] | [-4.09,3.23] |
| **Miscarriage/Stillbirth history** (ref: No) | | | | |
| Yes | 1.21 | | 1.41 | 1.16 |
| | [-1.17,3.59] | | [-1.13,3.95] | [-1.15,3.46] |
| **Number of ANC contacts** (ref: 1-3) | | | | |
| 0[§] | **7.33**$^{*}$ | | 7.10 | 5.24 |
| | **[0.57,14.09]** | | [-0.14,14.34] | [-1.25,11.72] |
| 4+ | **2.43**$^{*}$ | | 2.12 | 1.66 |
| | **[0.13,4.74]** | | [-0.25,4.50] | [-0.48,3.81] |
| **Place of ANC** (ref: Different facility/home/ no ANC) | | | | |
| Same facility as place of delivery | -0.20 | | 0.50 | 1.00 |
| | [-2.55,2.14] | | [-2.05,3.06] | [-1.34,3.34] |
| | | | - | - |
| | *HEATH SYSTEMS DETERMINANTS* | | | |
| **Delivery facility type** (ref: Health center) | | | | |
| Hospital | 3.40 | | | 0.41 |

| | | | | |
|---|---|---|---|---|
| | [-0.87,7.67] | | | [-3.94,4.77] |
| **Delivery facility managing authority/ownership** (ref: Public) | | | | |
| Private for profit | **8.99**$^{**}$ | | | **7.36**$^{*}$ |
| | **[3.20,14.77]** | | | **[1.29,13.44]** |
| Private non-profit/faith-based | **5.68**$^{**}$ | | | 3.01 |
| | **[1.68,9.69]** | | | [-1.39,7.42] |
| **Assistance during delivery** (ref: midwife/nurse/TBA) | | | | |
| Physician/specialist | -4.88 | | | -3.40 |
| | [-11.61,1.85] | | | [-9.62,2.82] |
| Other/unskilled§ | -0.81 | | | 11.27 |
| | [-15.45,13.83] | | | [-2.15,24.69] |
| Don't Know/Couldn't distinguish | **-3.35**$^{*}$ | | | -1.68 |
| | **[-6.03,-0.67]** | | | [-4.34,0.97] |
| **Maternal PNC before discharge** (ref: No) | | | | |
| Yes | **11.03**$^{**}$ | | | -1.35 |
| | **[7.74,14.33]** | | | [-6.58,3.89] |
| **Length of facility stay** (ref: <24h) | | | | |
| ≥24h | -1.26 | | | -1.80 |
| | [-3.72,1.20] | | | [-4.12,0.51] |
| **PNC counseling: danger signs** (ref: No) | | | | |
| Yes | **7.81**$^{**}$ | | | **2.99**$^{*}$ |
| | **[5.46,10.16]** | | | **[0.13,5.84]** |
| **PNC counselling: family planning** (ref: No) | | | | |
| Yes | **3.90**$^{**}$ | | | 0.27 |
| | **[1.62,6.18]** | | | [-2.06,2.60] |
| **PNC: BP check** (ref: No) | | | | |
| Yes | **8.54**$^{**}$ | | | **3.97**$^{*}$ |
| | **[5.96,11.12]** | | | **[0.61,7.34]** |
| **PNC: newborn check** (ref: No) | | | | |
| Yes | **13.07**$^{**}$ | | | **9.06**$^{**}$ |
| | **[9.92,16.21]** | | | **[4.74,13.38]** |
| **PNC: newborn appointment** (ref: No) | | | | |
| Yes | **8.28**$^{*}$ | | | 2.47 |
| | **[1.56,14.99]** | | | [-3.72,8.66] |
| Observations (n) | | 401 | 377 | 371 |
| AIC | | 3043.5 | 2883.0 | 2775.7 |

95% confidence intervals in brackets

$^{*}$ $p < 0.05$, $^{**}$ $p < 0.01$

§ small sample (≤10)

Supplemental tables 6A-C: Structural, intermediary and health systems determinants of PCMC domains by study site.

A. Lusaka

| | Dignity & Respect (out of 18 points) | Communication & Autonomy (out of 27 points) | Supportive Care (out of 45 points) |
|---|---|---|---|
| *STRUCTURAL DETERMINANTS* | | | |
| **Education** (ref: primary or less) | | | |
| secondary or more | -0.39<br>[-1.08,0.31] | 0.32<br>[-0.76,1.41] | -0.86<br>[-2.01,0.29] |
| **Employment** (ref: unemployed) | | | |
| Employed (public, private, self) | 0.03<br>[-0.65,0.71] | **1.15***<br>**[0.09,2.20]** | 0.16<br>[-0.95,1.27] |
| Informal/casual labor | -0.75<br>[-1.87,0.36] | 0.01<br>[-1.73,1.74] | -0.21<br>[-2.04,1.63] |
| *INTERMEDIARY DETERMINANTS* | | | |
| **Age** (ref: 20-35 yrs) | | | |
| 15-19 yrs | -0.56<br>[-1.65,0.54] | -1.35<br>[-3.05,0.35] | 0.33<br>[-1.47,2.13] |
| 35-49 yrs | 0.01<br>[-1.12,1.13] | -0.20<br>[-1.95,1.55] | -0.49<br>[-2.34,1.35] |
| **Marital status** (ref: in union) | | | |
| Not in union | -0.22<br>[-1.05,0.62] | 0.95<br>[-0.35,2.25] | 1.08<br>[-0.30,2.45] |
| **Parity** (ref: 2-3 children) | | | |
| 1 | 0.31<br>[-0.54,1.16] | 0.10<br>[-1.22,1.43] | -0.99<br>[-2.39,0.41] |
| 4+ | -0.30<br>[-1.20,0.61] | -0.10<br>[-1.51,1.31] | 0.18<br>[-1.30,1.67] |
| **Pregnancy complications** (ref: No) | | | |
| Yes | -0.17<br>[-1.24,0.90] | 0.74<br>[-0.92,2.41] | -1.45<br>[-3.21,0.31] |
| **Miscarriage/Stillbirth history** (ref: No) | | | |
| Yes | -0.26<br>[-1.09,0.56] | -0.23<br>[-1.52,1.06] | -0.19<br>[-1.56,1.17] |
| **Number of ANC contacts** (ref: 1-3) | | | |
| 0 | -1.61<br>[-4.25,1.03] | 1.00<br>[-3.12,5.11] | 0.29<br>[-4.05,4.63] |
| 4+ | 0.10<br>[-0.60,0.79] | 0.13<br>[-0.96,1.22] | 0.36<br>[-0.79,1.51] |
| **Place of ANC** (ref: Different facility/home/no ANC) | | | |
| Same facility as place of delivery | -0.39<br>[-1.06,0.28] | -0.53<br>[-1.58,0.52] | -0.58<br>[-1.69,0.53] |

| | | | |
|---|---|---|---|
| *HEALTH SYSTEMS DETERMINANTS* | | | |
| **Delivery facility type** (ref: Health center) | | | |
| Hospital | -0.11 | 0.24 | -1.34 |
| | [-1.44,1.22] | [-2.86,3.33] | [-3.94,1.26] |
| **Assistance during delivery** (ref: midwife/ nurse/TBA) | | | |
| Physician/specialist | 0.42 | **2.56**$^{**}$ | 1.07 |
| | [-0.62,1.46] | **[0.94,4.18]** | [-0.64,2.78] |
| Other/unskilled | -0.55 | 1.49 | -2.61 |
| | [-3.33,2.24] | [-2.86,5.83] | [-7.20,1.97] |
| Don't Know/Could not distinguish | -0.03 | -0.95 | -2.09 |
| | [-1.87,1.82] | [-3.82,1.93] | [-5.12,0.95] |
| **Maternal PNC before discharge** (ref: No) | | | |
| Yes | 1.72 | 0.59 | -0.55 |
| | [-0.45,3.89] | [-2.79,3.96] | [-4.11,3.02] |
| **Length of facility stay** (ref: <24h) | | | |
| ≥24h | -0.64 | -0.70 | -0.18 |
| | [-1.35,0.08] | [-1.81,0.40] | [-1.35,1.00] |
| **PNC counseling: danger signs** (ref: No) | | | |
| Yes | **1.12**$^{**}$ | **2.59**$^{**}$ | **3.02**$^{**}$ |
| | **[0.35,1.89]** | **[1.39,3.79]** | **[1.75,4.29]** |
| **PNC counseling: family planning** (ref: No) | | | |
| Yes | -0.34 | 0.03 | -0.58 |
| | [-1.05,0.37] | [-1.08,1.14] | [-1.75,0.59] |
| **PNC: BP check** (ref: No) | | | |
| Yes | 0.58 | -0.13 | 2.27 |
| | [-1.11,2.27] | [-2.75,2.50] | [-0.50,5.05] |
| **PNC: newborn check** (ref: No) | | | |
| Yes | 0.25 | 1.12 | **3.41**$^{**}$ |
| | [-1.10,1.59] | [-0.98,3.22] | **[1.20,5.63]** |
| **PNC: newborn appointment** (ref: No) | | | |
| Yes | 0.94 | -1.69 | 0.61 |
| | [-0.77,2.65] | [-4.36,0.97] | [-2.20,3.43] |
| Observations | 419 | 419 | 419 |

95% confidence intervals in brackets

$^{*}$ $p < 0.05$, $^{**}$ $p < 0.01$

B. Nairobi

| | Dignity & Respect (out of 18 points) | Communication & Autonomy (out of 27 points) | Supportive Care (out of 45 points) |
|---|---|---|---|
| *STRUCTURAL DETERMINANTS* | | | |
| **Education** (ref: primary or less) | | | |
| Secondary or more | -0.03 | 0.08 | -0.36 |
| | [-0.53,0.48] | [-0.79,0.95] | [-1.28,0.56] |
| **Employment** (ref: unemployed) | | | |
| Employed (public, private, self) | 0.52 | -0.37 | 0.25 |
| | [-0.04,1.07] | [-1.33,0.58] | [-0.76,1.26] |
| Informal/casual labor | 0.08 | -0.39 | 0.69 |
| | [-0.61,0.76] | [-1.57,0.79] | [-0.56,1.94] |
| *INTERMEDIARY DETERMINANTS* | | | |
| **Age** (ref: 20-35 yrs) | | | |
| 15-19 yrs | -0.22 | -0.31 | -0.25 |
| | [-1.21,0.78] | [-2.03,1.41] | [-2.08,1.57] |
| 35-49 yrs | **0.91*** | -0.25 | 1.20 |
| | **[0.14,1.68]** | [-1.58,1.08] | [-0.20,2.61] |
| **Marital status** (ref: in union) | | | |
| Not in union | -0.14 | **-1.24*** | -1.16 |
| | [-0.83,0.55] | **[-2.43,-0.05]** | [-2.42,0.11] |
| **Parity** (ref: 2-3 children) | | | |
| 1 | 0.00 | **-1.02*** | -0.26 |
| | [-0.59,0.60] | **[-2.03,-0.00]** | [-1.33,0.82] |
| 4+ | 0.24 | -0.28 | 0.65 |
| | [-0.53,1.01] | [-1.61,1.05] | [-0.76,2.06] |
| **Pregnancy complications** (ref: No) | | | |
| Yes | 0.00 | -0.29 | 1.00 |
| | [-0.65,0.66] | [-1.42,0.84] | [-0.20,2.20] |
| **Miscarriage/Stillbirth history** (ref: No) | | | |
| Yes | -0.37 | -0.30 | -0.60 |
| | [-1.00,0.26] | [-1.39,0.79] | [-1.76,0.56] |
| **Number of ANC contacts** (ref: 1-3) | | | |
| 0 | -1.65 | 0.70 | 0.25 |
| | [-3.98,0.69] | [-3.34,4.74] | [-4.03,4.53] |
| 4+ | 0.34 | 0.86 | 0.02 |
| | [-0.17,0.85] | [-0.02,1.74] | [-0.91,0.96] |
| **Place of ANC** (ref: Different facility/home/no ANC) | | | |
| Same facility as place of delivery | -0.22 | 0.14 | -0.32 |
| | [-0.75,0.32] | [-0.83,1.10] | [-1.35,0.71] |
| *HEALTH SYSTEMS DETERMINANTS* | | | |
| **Delivery facility type** (ref: Health center) | | | |
| Hospital | -0.33 | **-1.90*** | **-4.81**** |
| | [-0.98,0.32] | **[-3.63,-0.16]** | **[-7.11,-2.51]** |
| **Delivery facility managing authority/ownership** (ref: Public) | | | |
| Private for profit | **1.26*** | -0.45 | 1.67 |

| | | | |
|---|---|---|---|
| | **[0.27,2.26]** | [-2.69,1.79] | [-1.10,4.44] |
| Private non-profit/faith-based | **0.89**$^{*}$ | -0.58 | 0.67 |
| | **[0.10,1.67]** | [-2.45,1.28] | [-1.71,3.04] |
| **Assistance during delivery** (ref: midwife /nurse /TBA) | | | |
| Physician/specialist | **1.03**$^{**}$ | **2.03**$^{**}$ | **1.74**$^{**}$ |
| | **[0.55,1.52]** | **[1.16,2.90]** | **[0.81,2.66]** |
| Don't Know § | **2.16**$^{*}$ | 1.21 | -1.37 |
| | **[0.51,3.80]** | [-1.62,4.05] | [-4.37,1.63] |
| **Maternal PNC before discharge** (ref: No) | | | |
| Yes | 0.98 | 0.24 | 1.89 |
| | [-0.50,2.46] | [-2.32,2.81] | [-0.83,4.60] |
| **Length of facility stay** (ref: <24h) | | | |
| ≥24h | 0.38 | -0.73 | -0.48 |
| | [-0.26,1.02] | [-1.86,0.40] | [-1.68,0.71] |
| **PNC counseling: danger signs** (ref: No) | | | |
| Yes | **0.65**$^{*}$ | **2.88**$^{**}$ | **1.08**$^{*}$ |
| | **[0.12,1.18]** | **[1.94,3.81]** | **[0.09,2.08]** |
| **PNC counseling: family planning** (ref: No) | | | |
| Yes | -0.34 | -0.21 | -0.27 |
| | [-0.84,0.17] | [-1.14,0.71] | [-1.26,0.72] |
| **PNC: BP check** (ref: No) | | | |
| Yes | 0.27 | 0.95 | 1.02 |
| | [-0.62,1.15] | [-0.57,2.47] | [-0.59,2.63] |
| **PNC: newborn check** (ref: No) | | | |
| Yes | **1.51**$^{**}$ | **3.11**$^{**}$ | **2.84**$^{**}$ |
| | **[0.45,2.56]** | **[1.27,4.94]** | **[0.89,4.78]** |
| **PNC: newborn appointment** (ref: No) | | | |
| Yes | 0.41 | 0.51 | 1.05 |
| | [-0.87,1.69] | [-1.69,2.71] | [-1.28,3.38] |
| Observations | 405 | 405 | 405 |

95% confidence intervals in brackets

$^{*}$ $p < 0.05$, $^{**}$ $p < 0.01$

§ small sample (≤10)

C. Ouagadougou

| | Dignity & Respect (out of 18 points) | Communication & Autonomy (out of 27 points) | Supportive Care (out of 45 points) |
|---|---|---|---|
| *STRUCTURAL DETERMINANTS* | | | |
| **Education** (ref: primary or less) | | | |
| Secondary or more | -0.15 | 0.22 | 0.57 |
| | [-0.80,0.49] | [-0.71,1.15] | [-0.68,1.82] |
| **Employment** (ref: unemployed) | | | |
| Employed (public, private, self) | 0.19 | **1.94**** | 1.09 |
| | [-0.66,1.05] | **[0.70,3.17]** | [-0.57,2.74] |
| Informal/casual labor | 0.06 | 0.69 | 0.29 |
| | [-0.58,0.70] | [-0.23,1.61] | [-0.94,1.52] |
| *INTERMDIARY DETERMINANTS* | | | |
| **Age** (ref: 20-35 yrs) | | | |
| 15-19 yrs | 0.84 | 1.01 | 0.66 |
| | [-0.36,2.04] | [-0.72,2.74] | [-1.66,2.98] |
| 35-49 yrs | 0.34 | 0.33 | 0.84 |
| | [-0.61,1.29] | [-1.04,1.70] | [-0.99,2.68] |
| **Marital status** (ref: in union) | | | |
| Not in union | -0.86 | 0.29 | 1.11 |
| | [-3.13,1.41] | [-2.99,3.56] | [-3.27,5.49] |
| **Parity** (ref: 2-3 children) | | | |
| 1 | -0.43 | -0.36 | -0.42 |
| | [-1.24,0.38] | [-1.54,0.81] | [-1.99,1.15] |
| 4+ | -0.27 | 0.12 | -0.54 |
| | [-1.05,0.51] | [-1.01,1.24] | [-2.04,0.96] |
| **Pregnancy complications** (ref: No) | | | |
| Yes | -0.04 | 0.75 | -1.20 |
| | [-1.06,0.98] | [-0.74,2.23] | [-3.23,0.82] |
| **Miscarriage/Stillbirth history** (ref: No) | | | |
| Yes | 0.43 | -0.37 | 1.12 |
| | [-0.24,1.10] | [-1.33,0.60] | [-0.17,2.41] |
| **Number of ANC contacts** (ref: 1-3) | | | |
| 0[§] | 1.06 | **3.07*** | 1.05 |
| | [-0.81,2.94] | **[0.36,5.78]** | [-2.58,4.67] |
| 4+ | 0.35 | 0.66 | 0.62 |
| | [-0.27,0.98] | [-0.23,1.56] | [-0.57,1.82] |
| **Place of ANC** (ref: Different facility/home /no ANC) | | | |
| Same facility as place of delivery | 0.04 | 0.56 | 0.35 |
| | [-0.63,0.71] | [-0.42,1.53] | [-0.95,1.66] |
| *HEALTH SYSTEMS DETERMINANTS* | | | |
| **Delivery facility type** (ref: Health center) | | | |
| Hospital | -0.16 | 0.22 | 0.31 |
| | [-1.19,0.87] | [-1.35,1.79] | [-1.99,2.61] |
| **Delivery facility managing authority/ownership** (ref: Public) | | | |
| Private for profit | 0.84 | **2.80*** | **3.46*** |

| | | | |
|---|---|---|---|
| | [-0.63,2.31] | **[0.57,5.04]** | **[0.23,6.69]** |
| Private non-profit/faith-based | 0.23 | 0.63 | 2.24 |
| | [-0.82,1.28] | [-0.97,2.23] | [-0.09,4.57] |
| **Assistance during delivery** (ref: midwife/ nurse/ TBA) | | | |
| Physician/specialist | -0.54 | -2.01 | -0.72 |
| | [-2.33,1.26] | [-4.61,0.58] | [-4.19,2.75] |
| Other/unskilled § | **4.83*** | 1.90 | 4.81 |
| | **[0.94,8.71]** | [-3.70,7.51] | [-2.70,12.31] |
| Don't Know | -0.48 | -0.75 | -0.38 |
| | [-1.20,0.24] | [-1.81,0.32] | [-1.84,1.08] |
| **Maternal PNC before discharge** (ref: No) | | | |
| Yes | -0.39 | -1.91 | 1.35 |
| | [-1.91,1.12] | [-4.10,0.28] | [-1.58,4.28] |
| **Length of facility stay** (ref: <24h) | | | |
| ≥24h | -0.44 | -0.22 | -1.01 |
| | [-1.09,0.21] | [-1.17,0.73] | [-2.29,0.28] |
| **PNC counseling: danger signs** (ref: No) | | | |
| Yes | 0.68 | 0.22 | **2.05*** |
| | [-0.13,1.49] | [-0.96,1.40] | **[0.46,3.64]** |
| **PNC counseling: family planning** (ref: No) | | | |
| Yes | -0.19 | 0.68 | -0.27 |
| | [-0.85,0.47] | [-0.28,1.64] | [-1.56,1.03] |
| **PNC: BP check** (ref: No) | | | |
| Yes | 0.68 | **2.57**** | 0.68 |
| | [-0.29,1.65] | **[1.17,3.97]** | [-1.20,2.56] |
| **PNC: newborn check** (ref: No) | | | |
| Yes | **2.02**** | **3.57**** | **3.15*** |
| | **[0.79,3.25]** | **[1.78,5.36]** | **[0.74,5.56]** |
| **PNC: newborn appointment** (ref: No) | | | |
| Yes | **1.81*** | 2.16 | -1.31 |
| | **[0.01,3.60]** | [-0.43,4.76] | [-4.78,2.15] |
| Observations | 371 | 371 | 371 |

95% confidence intervals in brackets
* $p < 0.05$, ** $p < 0.01$
§ small sample (≤10)

Supplemental table 7 . PCMC supportive care item responses (% women) by delivery facility type in Nairobi (n=412)

| Question | Response category | Health center/other (n=195) (%) | Hospital (n=217) (%) | $Chi^2$ p value |
|---|---|---|---|---|
| How did you feel about the amount of time you waited to receive care? Would you say it was: | Very long | 51.8 | 30.4 | <0.001 |
| | Somewhat long | 33.3 | 39.2 | |
| | Somewhat short | 10.8 | 23.0 | |
| | Very short | 4.1 | 7.4 | |
| Did the doctors and nurses at the facility show concern for your feelings about your delivery? | No, never | 0.5 | 8.3 | <0.001 |
| | Yes, a few times | 7.7 | 31.8 | |
| | Yes, most of the time | 27.7 | 43.8 | |
| | Yes, all the time | 64.1 | 16.1 | |
| Did the doctors, nurses, or other staff at the facility try to understand your anxieties? | No, never | 4.1 | 12.0 | <0.001 |
| | Yes, a few times | 4.1 | 21.2 | |
| | Yes, most of the time | 15.9 | 13.4 | |
| | Yes, all the time | 21.0 | 6.5 | |
| | Not applicable | 54.9 | 47 | |
| When you needed help, did you feel the doctors, nurses, or other staff at the facility paid attention? | No,never | 2.1 | 3.2 | <0.001 |
| | Yes, a few times | 7.7 | 25.8 | |
| | Yes, most of the time | 27.2 | 50.2 | |
| | Yes, all the time | 63.1 | 20.7 | |
| Do you feel the doctors or nurses did everything they could to help control your pain? | No,never | 17.9 | 19.4 | <0.001 |
| | Yes, a few times | 10.3 | 24.4 | |
| | Yes, most of the time | 22.1 | 35.5 | |
| | Yes, all the time | 49.7 | 20.7 | |

| | | | | |
|---|---|---|---|---|
| Were you allowed to have someone you wanted (outside of staff at the facility, such as family or friends) to stay with you during labor? | No, never | 67.2 | 84.8 | <0.001 |
| | Yes, a few times | 17.9 | 7.8 | |
| | Yes, most of the time | 2.1 | 2.3 | |
| | Yes, all the time | 8.2 | 0.9 | |
| | I did not want someone to stay with me | 4.6 | 4.1 | |
| Were you allowed to have someone you wanted to stay with you during delivery? | No, never | 86.2 | 90.3 | 0.305 |
| | Yes, a few times | 1.5 | 1.8 | |
| | Yes, most of the time | 0.5 | 0.5 | |
| | Yes, all the time | 3.1 | 0.5 | |
| | I did not want someone to stay with me | 8.7 | 6.9 | |
| Did you feel the doctors, nurses, or other staff at the facility took good care of you, at the best of their ability? | No,never | 0.5 | 2.8 | <0.001 |
| | Yes, a few times | 1.5 | 8.8 | |
| | Yes, most of the time | 24.6 | 56.2 | |
| | Yes, all the time | 73.3 | 32.3 | |
| Did you feel you could completely trust the doctors, nurses, or other staff at the facility with regards to your care? | No,never | 1 | 0.9 | <0.001 |
| | Yes, a few times | 3.1 | 6.5 | |
| | Yes, most of the time | 21.5 | 64.1 | |
| | Yes, all the time | 74.4 | 28.6 | |
| Do you think there were enough health staff in the facility to care for you? | No,never | 2.1 | 0.0 | <0.001 |
| | Yes, a few times | 6.2 | 17.1 | |
| | Yes, most of the time | 19.5 | 50.2 | |
| | Yes, all the time | 72.3 | 32.7 | |

| | | | | |
|---|---|---|---|---|
| Thinking about the labor and postnatal wards, did you feel the health facility was crowded? | Yes, all the time | 6.2 | 29.5 | <0.001 |
| | Yes, most of the time | 3.6 | 11.5 | |
| | Yes, a few times | 9.2 | 14.7 | |
| | No,never | 81.0 | 44.2 | |
| Thinking about the wards, washrooms, and the general environment of the health facility, would you say the facility was very clean, clean, dirty, or very dirty? | Very dirty | 0.0 | 0.0 | <0.001 |
| | Dirty | 0.0 | 5.5 | |
| | Clean | 23.6 | 69.1 | |
| | Very clean | 76.4 | 25.3 | |
| Was there running water in the facility? | No,never | 1.5 | 0.5 | 0.002 |
| | Yes, a few times | 2.1 | 1.4 | |
| | Yes, most of the time | 4.1 | 15.2 | |
| | Yes, all the time | 92.3 | 82.9 | |
| Was there electricity in the facility? | No,never | 0.5 | 0.0 | 0.005 |
| | Yes, a few times | 0.5 | 0.0 | |
| | Yes, most of the time | 3.6 | 12.4 | |
| | Yes, all the time | 95.4 | 87.6 | |
| In general, did you feel safe in the health facility? | No,never | 0.0 | 0.9 | <0.001 |
| | Yes, a few times | 0.5 | 3.2 | |
| | Yes, most of the time | 6.2 | 47.5 | |
| | Yes, all the time | 93.3 | 48.4 | |
